# Unveiling the crucial neuronal role of the proteasomal ATPase subunit gene *PSMC5* in neurodevelopmental proteasomopathies

**DOI:** 10.1101/2024.01.13.24301174

**Authors:** Sébastien Küry, Janelle E. Stanton, Geeske van Woerden, Tzung-Chien Hsieh, Cory Rosenfelt, Marie Pier Scott-Boyer, Victoria Most, Tianyun Wang, Jonas Johannes Papendorf, Charlotte de Konink, Wallid Deb, Virginie Vignard, Maja Studencka-Turski, Thomas Besnard, Anna Marta Hajdukowicz, Franziska Thiel, Sophie Möller, Laëtitia Florenceau, Silvestre Cuinat, Sylvain Marsac, Ingrid Wentzensen, Annabelle Tuttle, Cara Forster, Johanna Striesow, Richard Golnik, Damara Ortiz, Laura Jenkins, Jill A. Rosenfeld, Alban Ziegler, Clara Houdayer, Dominique Bonneau, Erin Torti, Amber Begtrup, Kristin G. Monaghan, Sureni V. Mullegama, C.M.L. (Nienke) Volker-Touw, Koen L. I. van Gassen, Renske Oegema, Mirjam de Pagter, Katharina Steindl, Anita Rauch, Ivan Ivanovski, Kimberly McDonald, Emily Boothe, Andrew Dauber, Janice Baker, Noelle Andrea V Fabie, Raphael A. Bernier, Tychele N. Turner, Siddharth Srivastava, Kira A. Dies, Lindsay Swanson, Carrie Costin, Rebekah K. Jobling, John Pappas, Rachel Rabin, Dmitriy Niyazov, Anne Chun-Hui Tsai, Karen Kovak, David B. Beck, MCV Malicdan, David R. Adams, Lynne Wolfe, Rebecca D. Ganetzky, Colleen Muraresku, Davit Babikyan, Zdeněk Sedláček, Miroslava Hančárová, Andrew T. Timberlake, Hind Al Saif, Berkley Nestler, Kayla King, MJ Hajianpour, Gregory Costain, D’Arcy Prendergast, Chumei Li, David Geneviève, Antonio Vitobello, Arthur Sorlin, Christophe Philippe, Tamar Harel, Ori Toker, Ataf Sabir, Derek Lim, Mark Hamilton, Lisa Bryson, Elaine Cleary, Sacha Weber, Trevor L. Hoffman, Anna Maria Cueto-González, Eduardo Fidel Tizzano, David Gómez-Andrés, Marta Codina-Solà, Athina Ververi, Efterpi Pavlidou, Alexandros Lambropoulos, Kyriakos Garganis, Marlène Rio, Jonathan Levy, Sarah Jurgensmeyer, Anne M. McRae, Mathieu Kent Lessard, Maria Daniela D’Agostino, Isabelle De Bie, Meret Wegler, Rami Abou Jamra, Susanne B. Kamphausen, Viktoria Bothe, Larissa M. Busch, Uwe Völker, Elke Hammer, Kristian Wende, Benjamin Cogné, Bertrand Isidor, Jens Meiler, Amélie Bosc-Rosati, Julien Marcoux, Marie-Pierre Bousquet, Jeremie Poschmann, Frédéric Laumonnier, Peter W. Hildebrand, Evan E. Eichler, Kirsty McWalter, Peter M. Krawitz, Arnaud Droit, Ype Elgersma, Andreas M. Grabrucker, Francois V. Bolduc, Stéphane Bézieau, Frédéric Ebstein, Elke Krüger

## Abstract

Neurodevelopmental proteasomopathies represent a distinctive category of neurodevelopmental disorders (NDD) characterized by genetic variations within the 26S proteasome, a protein complex governing eukaryotic cellular protein homeostasis. In our comprehensive study, we identified 23 unique variants in *PSMC5*, which encodes the AAA-ATPase proteasome subunit PSMC5/Rpt6, causing syndromic NDD in 38 unrelated individuals. Overexpression of *PSMC5* variants altered human hippocampal neuron morphology, while *PSMC5* knockdown led to impaired reversal learning in flies and loss of excitatory synapses in rat hippocampal neurons. *PSMC5* loss-of-function resulted in abnormal protein aggregation, profoundly impacting innate immune signaling, mitophagy rates, and lipid metabolism in affected individuals. Importantly, targeting key components of the integrated stress response, such as PKR and GCN2 kinases, ameliorated immune dysregulations in cells from affected individuals. These findings significantly advance our understanding of the molecular mechanisms underlying neurodevelopmental proteasomopathies, provide links to research in neurodegenerative diseases, and open up potential therapeutic avenues.

## Introduction

The proper structure and function of eukaryotic cells rely on the maintenance of the intracellular proteome composed of thousands of proteins. The dynamic balance between protein synthesis and degradation is orchestrated by the proteostatic network^1,2^, an integral component of which is the ubiquitin-proteasome system (UPS), that selectively eliminates short- and long-lived proteins, as well as misfolded proteins modified with ubiquitin^3^. At the very heart of the UPS is the 26S proteasome, a large multi-subunit protease consisting of a 20S core particle and a 19S regulatory particle, comprising a lid and a base^3^. The lid subunits recognize and bind polyubiquitinated target proteins^1^, while the base subunits Rpt1-6/PSMC1-6 utilize ATP hydrolysis to deubiquitinate, unfold and translocate substrates into the proteolytic chamber of the 20S core particle, where they are degraded into oligopeptides. These oligopeptides can be further hydrolyzed into amino acids by peptidases, enabling efficient recycling within the cellular protein synthesis machinery^1,3^.

The remarkable dynamics and plasticity of proteasomes impart a significant impact on cellular physiology^3^, particularly in neurons. Neuronal synaptic plasticity relies on continuous proteome renewal, necessitating robust translation at ribosomes and concurrent breakdown of defective ribosomal products^2,4^. This maintenance of protein homeostasis is particularly challenging in cortical neurons, where synapses may contain hundreds of proteins^4^. Effective protein clearance by the proteasome is crucial for various neuronal processes, including synaptic remodeling, cell migration, neurotransmitter release, long-term potentiation, long-term depression, and memory formation^5,6^. Inhibition of proteasome activity has been experimentally shown to disrupt synapse composition, promote the aggregation of polyubiquitinated proteins, and ultimately lead to neurodegeneration^6^. Similar observations have been made in brain tissues from individuals with neurodegenerative disorders^6^ and schizophrenia^7^. Genetic investigations have further unveiled that loss-of-function variants in proteasome genes contribute to the onset and progression of a unique class of NDDs, specifically referred to as the neurodevelopmental proteasomopathies^8^. The inheritance pattern for these disorders can either be dominant, as seen with *PSMD12* variants associated with Stankiewicz-Isidor syndrome [MIM: 617516]^9,10^ and *PSMC3*^11^, or recessive, as observed in cases involving *PSMB1* [MIM: 620038]^12^ and *PSMC3* [MIM:619354]^13^ variants.

In this context, the proteasome subunit PSMC5/Rpt6 is of particular interest in this context due to its role in regulating proteasome activity and synaptic remodeling. As an ATPase, PSMC5 serves as a neuronal sensor that dynamically adjusts proteasome activity and localization in response to protein turnover requirements through phosphorylation by CAMKIIα^14^. The present study further confirms the pivotal role of *PSMC5* in neuronal function. We describe 23 variants in *PSMC5*, the vast majority heterozygous and *de novo*, in 38 unrelated individuals with delayed neurodevelopment. Functional analyses performed on patient cells and animal models unveiled that *PSMC5* alterations lead to a loss of proteasome function, triggering a complex cellular program involving profound remodeling of innate immunity, inflammation, and lipid metabolism. Intriguingly, the integrated stress response (ISR) mediates most of these changes, and potential molecular targets have been identified, holding promise for the development of biomarkers and therapeutic interventions. This study offers critical insights into the multifaceted impact of PSMC5 variants on NDD, advancing our understanding of the intricate cellular mechanisms underlying these conditions.

## Materials and Methods

### Genetic studies and ethics statement

The 38 affected individuals of the present study were enrolled =together with their healthy parents whenever possible= by participating teams involved in diagnostic or research activities on developmental disorders. All 38 affected individuals were clinically assessed by at least one clinical geneticist from each participating center, and gave written informed consent for participating in the study. Genome- or exome sequencing was performed in all affected individuals, and most often on their parents too, using a trio approach. Connections between the participating centers were aided by the web-based tool GeneMatcher. Consent for the publication of photographs was obtained for subjects S1, S3, S9, S10, S12, S13, S19, S30-34 and S38. The photographs are not shown in the current version of the manuscript but are deposited in GestaltMatcher database; accession links are provided in Figure 2, in place of the original pictures. The study has been approved by the CHU de Nantes-ethics committee (number CCTIRS: 14.556). This research was ethically approved by CPP Ouest V (File 06/15) on 04/08/2015 (Ref MESR DC 2017 2987).

**Figure 1:**
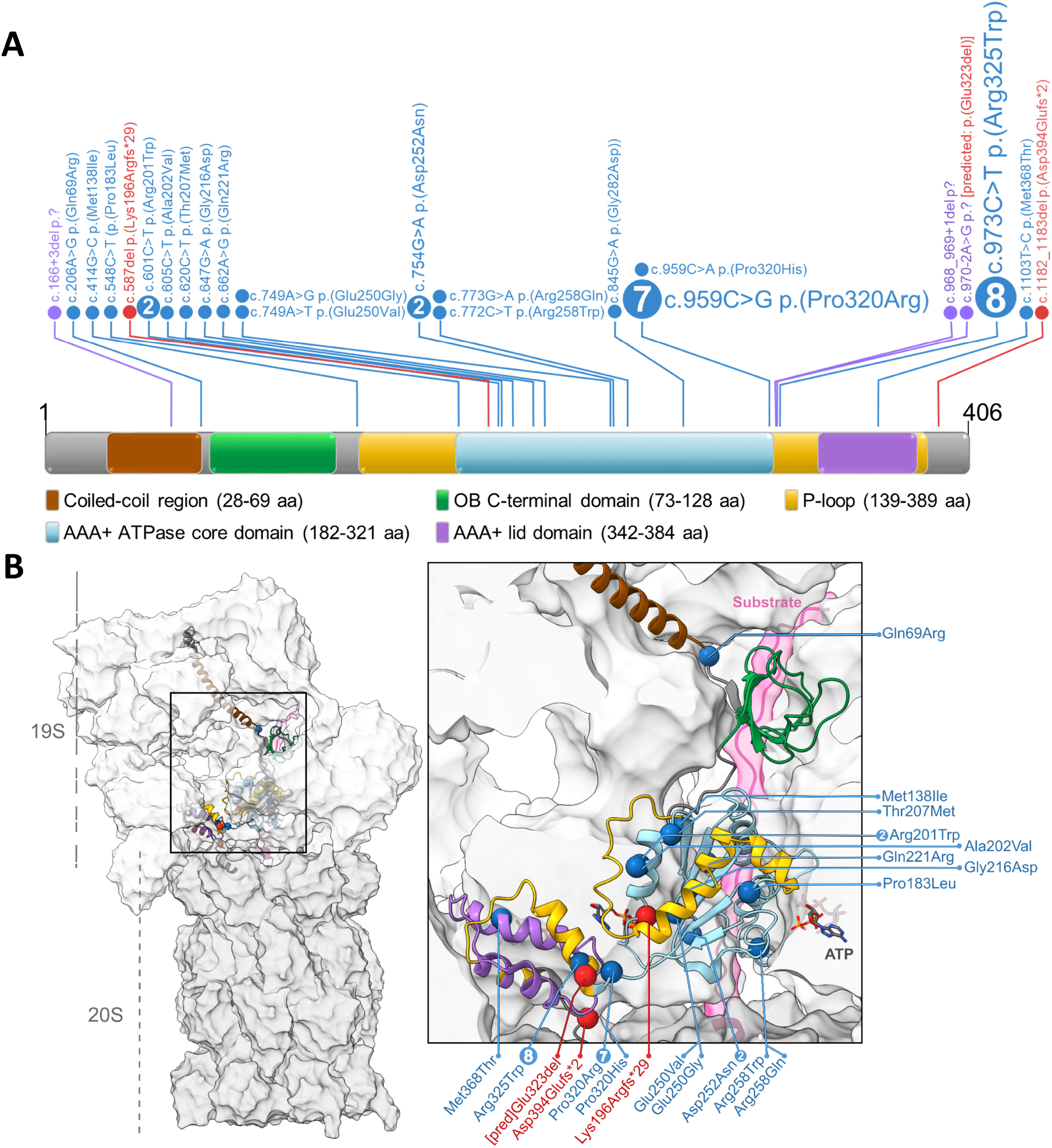
Structural analysis of *PSMC5* variants reveals most missense variants are located in the AAA+ ATPase domain of PSMC5/Rpt6 and affect proteasome dynamics or subunit interactions. *A*. Schematic representation of the PSMC5 (i.e. RPT6) protein on which the 23 alterations identified in patients with NDD are designated. Approximately half of these mutations fall into three distinct hotspot regions within the AAA+ ATPase domain of PSMC5/RPT6, as indicated. Numbers refer to the sums of unrelated NDD subjects in which the highlighted variant has been identified. Missense variants are colored in blue, frameshift variants or indels in red and splice site variants in purple. *B*. Based on the structures 6MSK from RCSB Protein Data Bank, we localized the missense variants identified in the AAA+ ATPase subunit PSMC5/Rpt6 within the 26S proteasomal complex. The variants of interest (spheres) are primarily located in the lower ATPase domain. Part of the 26S proteasome was hidden for better visibility; Rpt6 is shown in cartoon representation.

**Figure 2:**
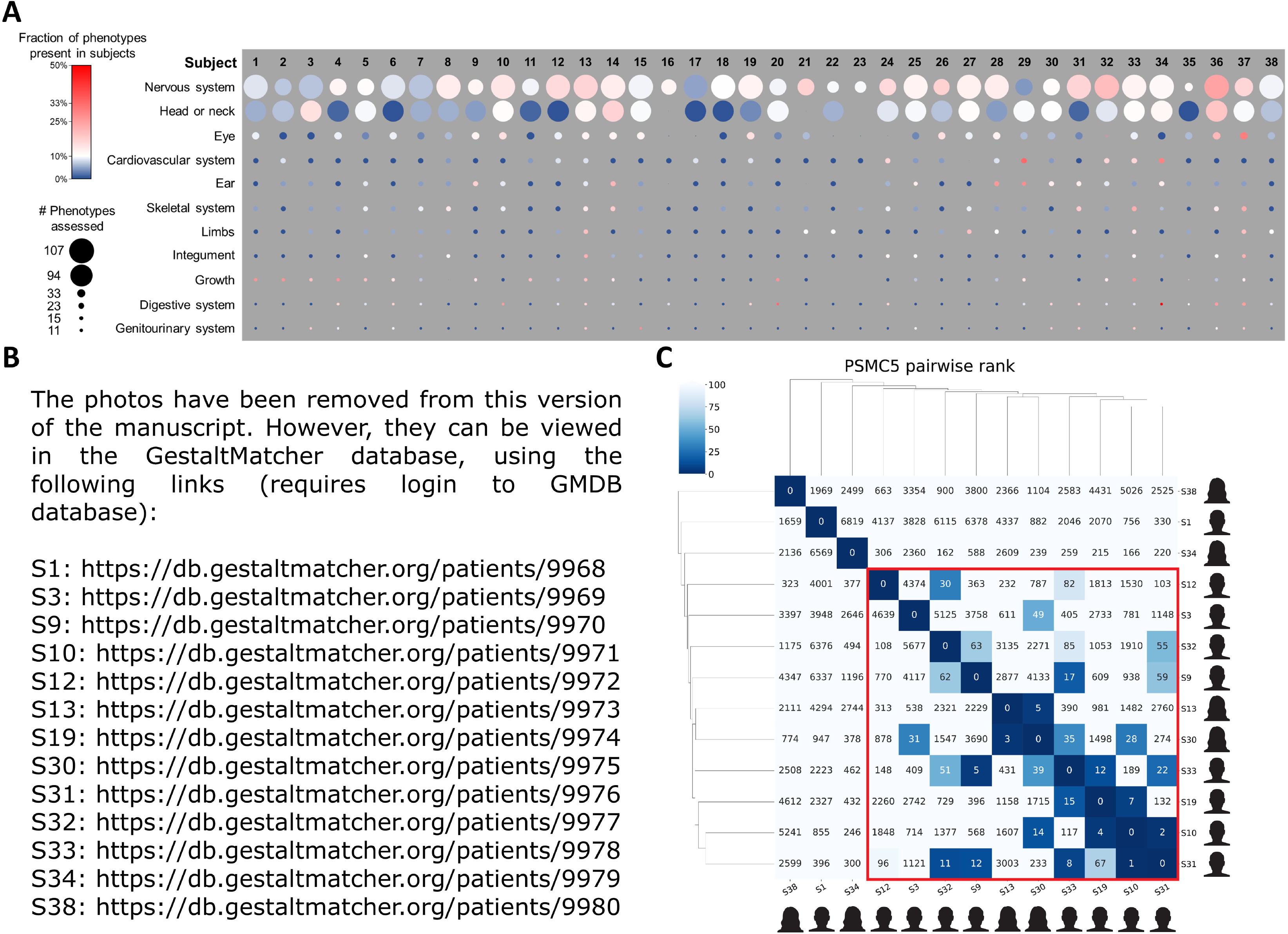
Clinical phenotypes associated with *PSMC5* variants. ***A.*** To visually represent the phenotypic data from tables S2b-2c, we organized the HPO terms into categories and ranked the top categories based on their prevalence within the cohort. Within each category, circles illustrate the number of phenotypes assessed and documented for each individual. The size of a circle corresponds to the number of phenotypes evaluated for a particular subject. If no phenotypes were assessed, there is no circle present. The color of each circle indicates the proportion of these confirmed phenotypes in a subject, with a spectrum ranging from dark blue (indicating the lowest fraction) to red (indicating the highest fraction). The top categories are arranged in descending order according to the highest average fraction observed among subjects. Notably, the most commonly observed categories among these are nervous system and head and neck anomalies. Intriguingly, there was no apparent correlation between genotype and phenotype, as significant variability in phenotypic expression was observed even among individuals with the same genetic variant. ***B*.** Dysmorphic facial features included notably abnormal ear morphology as seen in subjects S3/9/10/13/19/30/32-34 (this feature is present in 19/31 (61%) subjects in the whole cohort), abnormal palpebral fissures (13/35; 37% in the whole cohort) including downslanted ones as in subjects S3/10/13/30, thin upper lip vermilion as in subjects S3/34 (7/35; 20%), abnormal palate as in subject S10 (7/35; 20%), tall or broad forehead as in S3/33/34 (9/35; 26%), epicanthus as in S4/38 (5/35; 14%), and orofacial clefts as in subject S10 (2/35; 6%). ***C.*** Facial image analysis using GestaltMatcher. The pairwise rank matrix and hierarchical clustering of 13 *PSMC5* subjects. Each column is the result of testing one subject in the column and ranking of the remaining 12 photos in each row. For example, by testing the similarities between S10 and the 7,459 images of affected individuals from GMDB, S31 was ranked 1st and S19 was ranked 7th as being most similar to S10. The red box is the cluster of subjects with at least one match below the rank 50, which indicates this cluster shares a similar facial phenotype.

### Three-dimensional (3D) structural analysis of the effect of the *PSMC5* missense variants

This analysis was carried out to investigate the spatial localization within PSMC5/Rpt6 of the amino acid substitutions corresponding to the missense variants. Our objective was to offer insights into potential effects of the mutations on subunit arrangements, interactions, and related conformational changes. To achieve this, we employed two distinct structures: a Cryo-EM structure engaged with a substrate (PDB ID: 6MSK) and a substrate-free structure (PDB ID: 7W37). These structures allowed us to place the AAA+ ATPase subunit PSMC5/Rpt6 in the context of the 26S proteasome, specifically within the 19S regulatory particle. The resulting refined 3D map was then visualized using molecular visualization software UCSF ChimeraX.

### Facial image analysis

We performed the GestaltMatcher approach^15^ on *PSMC5* subjects to analyze the facial similarities among the 13 subjects (S1, S3, S9, S10, S12, S13, S19, S30-34 and S38) whose parents consented to facial analysis. We first utilized the model ensemble and test-time augmentation to encode each photo into 12 512-dimensional vectors. Each image *i* was represented in the space as 12 512-dimensional vectors x_i,k_ E lff and k E [1, 12]. The similarity between two subjects can be quantified by averaging 12 cosine distances between two subjects. When the cosine distance is smaller, the two subjects are more similar. To validate whether the subjects of the given cohort C, *PSMC5* subjects, are similar to each other, we performed the statistical analysis at the cohort level and the pairwise comparison analysis at the individual level.

We first performed a statistical analysis based on the mean pairwise cosine distance. We selected 1,555 images from different subjects with 328 different syndromes from GestaltMatcher Database (GMDB)^16^ and sampled two control distributions: (1) distances between subjects of the same syndrome and (2) distances between random patients. In the end, we compared the mean pairwise distance between patients of the given cohort C to these two distributions.

For each syndrome *S* and a random cohort size *n* (2< n < |S|), we sampled two cohorts: (1) C_S_ consisting of *n* patients from the chosen syndrome *S*, (2) C_R_ consisting of *n* randomly chosen patients from GMDB. We then calculated the mean pairwise cosine distance for each of two cohorts. This was repeated 100 times per syndrome, duplicated sampled cohorts were removed, resulting in a distribution of mean pairwise cosine distances between (1) patients stemming from the same syndrome, and (2) random patients, respectively.

We conducted a Receiver Operating Characteristic (ROC) analysis on the sampled cohorts from the same syndrome (“controls”) and from random patients (“cases”). We used the mean pairwise cosine distance as a measure of discrimination and performed the 5-fold cross-validation. We chose the threshold *c* by selecting the fold with the highest Youden index to decide whether the patients inside a given cohort are similar (mean pairwise cosine distance < *c*) or different (mean pairwise cosine distance > *c*), resulting in *c=0.915*, corresponding to a sensitivity of 0.851, a specificity of 0.862, and an Area Under the Curve (AUC) of 0.895

To examine the similarity of the *PSMC5* subjects, we calculated their mean pairwise cosine distances and compared them to the sampled distributions originating from patients from the same syndrome and random patients. Then, we sampled a cohort C_C_ of random sample size *n* (2<n < |C|) consisting of *n* patients from the given cohort C and calculated the mean pairwise cosine distance of this cohort. This was repeated 10,000 times and duplicated sampled cohorts were removed resulting in a distribution of mean pairwise cosine distances between patients stemming from the given cohort C.

In the end, we performed the pairwise comparison analysis on 13 *PSMC5* subjects. To simulate the real-life scenario, we first compared photographs of the 13 *PSMC5* subjects to 7,459 images with 449 different disorders stored in GMDB. For each *PSMC5* subject tested, we performed a leave-one-out cross-validation by placing the other 12 *PSMC5* subjects into the space of the 7,459 images and calculating the ranks of 12 subjects relative to the *PSMC5* subject tested.

### Behavioral studies in fly

#### Background

Rpt6 is the Drosophila ortholog of PSMC5. When using the DRSC integrative ortholog prediction tool (DIOPT), it has a rank of high homology and a weighted score of 15/16. The KD approach was substantiated by the presence of truncating variants in the cohort (V5:p.(Lys196Argfs*29) and V23:p.(Asp394Glufs*2)) and the resultant loss of proteasomal function.

#### Drosophila Strains

The *Rpt6* (*Psmc5)* Drosophila RNAi stock (34712) was obtained from Bloomington Drosophila Stock Center. Wild-type and ELAVGAL4 stocks were obtained from Dr. Tim Tully (Cold Spring Harbor Laboratory). Stocks were raised on a cornmeal agar medium at 22°C and 40% humidity. University of Alberta ethics and regulations were followed.

#### Genetic Crosses

Virgin females of wild-type or ELAVGAL4 were collected and crossed to theUAS-RNAi males.

#### Olfactory Learning

Drosophila olfactory learning was conducted by training 100 flies to associate an odour with a simultaneously presented footshock. A second control odour was then presented without a footshock. Flies were then funneled into a choice point and given a choice between the two odours. Flies that had correctly learned the association would avoid the odour that had been paired with the shock. This entire process was then repeated with 100 naive flies but with the odour shock pairing reversed in order to rule out any odour bias. The combined performance between these two trials constituted the performance index and n=1.

In reversal olfactory learning, the same procedure as normal learning was conducted with the added step of an additional training cycle after, in which the stimulus pairing was reversed. For example, flies were presented with odour A paired with a footshock and then odour B without footshock for the initial training cycle, followed by an additional training cycle where now odour B was paired with a footshock, and odour A was presented without. This is a more challenging task to perform as flies are required to unlearn the initial association in addition to learning the new pairing. Once again, this entire process was then repeated with 100 naive flies but with the odour pairings reversed in order to rule out any odour bias. Experiments were conducted at 25°C and 70% humidity. Adult Drosophila were transferred to bottles the day before testing. At the time of testing, flies were ages1-3 days.

#### Statistical analysis

For the learning experiments, a two-tailed t-test was performed. For the reversal learning experiments, a one-way ANOVA followed by Tukey’s test was performed.Prism software was used for all statistical analysis.

### Analysis of *PSMC5* knockdown in rat hippocampal neurons

#### Cultivation of primary neurons

Rat hippocampal neurons (RN-h, P10101, Innoprot) were plated on poly-L-lysine (PLL) (0.1 mg/mL)-coated coverslips in a 24-well plate at a density of 5×10^4^ on coverslips placed in a 24-well plate. RN-h were cultivated using Neurobasal medium containing 2% B27 (#17504044, Gibco), 1% Glutamax (#35050079, Gibco), 100 μg/mL penicillin and streptomycin. Cultures were incubated at 37°C with 5% CO_2_ in a sterile CO_2_ incubator for 14 days.

#### Lentiparticle-based Psmc5 knockdown

24 hours after plating, *Psmc5* - Rat shRNA lentiviral particles (TL711271V, Origene) were introduced into the culture medium at a multiplicity of Infection rate of 5 and incubated for 18 hours before a complete medium change was carried out.

#### Immunocytochemistry

Neurons were first washed with 1x PBS and fixed using 4% PFA for 15 min at room temperature. Permeabilization was performed by applying 0.2% Triton-X for 5 min and washing with 1x PBS. Next, blocking solutions (BS) (10% FBS/ 1x PBS) was applied for 1 h at room temperature. After that, the primary antibody (MAP2 (1:1000, Synaptic Systems #18006); vGlut (1:1000, Synaptic Systems #135304); vGAT (1:200, Synaptic System #131003)); PSMC5 (1:400, Sigma-Aldrich)) in BS was applied, and cells were incubated at 4°C overnight. Next, 1x PBS was applied for 3x 5 min, followed by incubating the secondary antibody diluted in BS in darkness for 1 h at room temperature. Cells were washed with 1x PBS twice for 5 min. An additional wash step includes the addition of 4,6-di-amino-2-phenylindol (DAPI) diluted in 1xPBS for 5 min. Finally, the cells were washed with 1x PBS, and a wash step with ddH_2_O was also carried out for 5 min. An ImageXpress Micro Confocal microscope (Molecular Devices, San Jose, USA) was used to obtain the images at a 40X magnification. Image analysis was carried out using ImageJ v1.53.

### Analysis of neuronal morphology in mouse

#### Cloning

Constructs containing human *PSMC5*^WT^ or mutated *PSMC5* cDNA sequences were isolated from the pcDNA3.1/Zeo(+) expression vector by PCR and tagged with AscI and PacI sequences, to allow ligation in the dual promotor expression vector, containing a CAGG-promotor for expression of the gene of interest and a PGK-promotor driven tdTomato gene to easily distinguish transfected cells from non-transfected cells. The following primers were used: (AscI) FW 5’-gaatccggcgcgccaccatggcgcttgacggaccagagc and (PacI) RV 5’-gaatccttaattaatcacttccataatttcttgatg. The empty vector control (EV) in the *in vitro* assays is merely the backbone plasmid, lacking any *PSMC5* sequence.

#### Mice

For the primary hippocampal neurons, FvB/NHanHsd females (ordered at 6-8 weeks old from Envigo) were crossed with FvB/NHanHsd females (ordered at 6-8 weeks old from Envigo). All mice were group-housed in IVC cages (Sealsage 1145T, Tecniplast) with bedding material (Lignocel BK 8/15 from Rettenmayer) on a 12/12 h light/dark cycle in 21°C (±1°C), humidity at 40-70% and with food pellets (801727CRM(P) from Special Dietary Service) and water available *ad libitum*. All animal experiments were conducted in accordance with the European Commission Council Directive 2010/63/EU (CCD project license AVD101002017893), and all described experiments and protocols were subjected to ethical review (and approved) by an independent review board (IRB) of the Erasmus MC.

#### Primary hippocampal cultures

To obtain primary hippocampal neurons, hippocampi were isolated from brains from E16.5 embryos and collected in ice cold Neural Basal Medium (NB; Gibco, 21103049). After two washes with ice cold NB, the hippocampi were incubated with Trypsin/EDTA (Sigma; T3924) for 20 min at 37°C. The tissue was again washed twice with warm NB and finally kept in supplemented NB (1%penicillin/streptomycin (Sigma; p4333)/1%GlutaMax (Gibco; 350500-38)/2%B27-supplement (Gibco; 17504044) for dissociation. Single cells were seeded in 12-wells plates containing Poly-D-Lysine (Sigma; P0899) coated coverslips and 1ml supplemented NB per well. The cells were incubated at 37°C/5% CO_2_ until transfection.

#### Neuronal transfections and immunological stainings

Primary hippocampal neurons (e16.5) were transfected on DIV3 with the following constructs: Empty expression vector; *PSMC5*^WT^ having the wild type human cDNA sequence referring to mRNA sequence NM_002805.6; PSMC5^Arg201Trp^ containing variant c.601C>T p.(Arg201Trp); *PSMC5*^Ala202Val^ containing variant c.605C>T p.(Ala202Val); *PSMC5*^Thr207Met^ containing variant c.620C>T p.(Thr207Met); *PSMC5*^Pro320His^ containing variant c.959C>A p.(Pro320His); *PSMC5*^Pro320Arg^ containing variant c.959C>G p.(Pro320Arg); *PSMC5*^Arg325Trp^ containing variant c.973C>T p.(Arg325Trp); *PSMC5*^Δex10^ having a deletion of exon 10 mimicking the likely consequence of splice site variant V20:c.970-2A>G. 2.5μg DNA was transfected of each PSMC5 construct. As control, 1.8μg DNA of the empty expression vector was transfected. Neurons were transfected by mixing the DNA with NB and Lipofectamin2000 according to the manufacturer’s instructions (Invitrogen; 11668-019)

The cells were fixed 5 days post-transfection (DIV8) using 4%PFA/4%sucrose. Fixed cells were labeled for MAP2 (1:500, Synaptic System; #188004) by o/n incubation at 4°C and visualized with a conjugated secondary antibody donkey-anti-guinea pig Alexa647 (1:200; Jackson ImmunoResearch #706-605-148), incubated for 1h at RT and finally covered with Mowiol (home-made) to allow for fluorescence imaging using confocal microscopy.

#### Confocal microscopy

All images were acquired by using a LSM700 confocal microscope (Zeiss). For imaging overexpression in primary hippocampal neurons, images were taken from 10 transfected neurons per condition, per neuronal batch (20x objective, 0.5 zoom, 2048 x 2048).

#### Analysis and statistics

The total neurite length and number of branches was traced using the program ImageJ and its plug-in NeuronJ. Within every batch, the data was normalized to the PSMC5^WT^, allowing us to pool the normalized data from different batches. For statistics, a one-way ANOVA (Dunnett’s multiple comparison test) was done, using Prism GraphPad. The n represents data from an individually traced neuron. For every condition a minimum of 8 neurons were imaged per batch, for a minimum of 2 independent primary hippocampal neuron batches.

### Expansion of T cells from PBMC isolated from control and patient samples

Whole-blood samples collected from healthy individuals as well as Subjects S1, S6, S11, S12, S21, S32 and S33 and their respective parents and/or siblings =whenever possible= were processed using spin medium gradient centrifugations (pluriSelect) to separate and isolate peripheral blood mononuclear cells (PBMC) for cryopreservation using a standard freezing medium consisting of 90% FBS and 10 % DMSO. At a later point in time, PBMC were plated on 96-well plates together with irradiated allogeneic PBMC in the presence of IL-2 and PHA-L for T cell expansion as in ^11^. Human T cells were maintained in RPMI1640 supplemented with 10 % human AB serum and 1% penicillin/streptomycin and analyzed at a resting state after 3-4 weeks of expansion.

### SDS-PAGE and western-blot analysis

Resting T cells and/or SH-SY5Y neuroblastoma cells were lysed in RIPA buffer (50 mM Tris pH 7.5, 150 mM NaCl, 2 mM EDTA, 1% NP40, 0.1% SDS) and proteins were quantified using a standard BCA assay (Thermofisher). Twenty micrograms of protein lysates were loaded on 10-15% SDS-PAGE and subsequently transferred onto PVDF membranes under wet conditions (200V, 400 mA, 1 h at 4°C). Following a 20-min blocking with ROTI®Block (Carl Roth), membranes were probed with primary antibodies overnight at 4°C under permanent shaking. Antibodies used in this study were directed against HA (BioLegend, clone HA.11), PSMC5/Rpt6 (Enzo Life Sciences, clone p45-110), α-tubulin (Abcam, clone DM1A), α6 (Enzo Life Sciences, clone MCP20), ubiquitin K48-linked ubiquitin chains (Cell Signaling, clone D9D5), GAPDH (Cell Signaling, clone 14C10), GRP94 (Invitrogen, PA5-27860), ATF6 (Cell Signaling, clone D4Z8V), BNIP3L (Cell Signaling, 12396). Antibody binding was detected using HRP-conjugated anti-mouse, -rabbit or -goat secondary antibodies and chemiluminescence (Biorad) following the manufacturer’s recommendations.

### RNA isolation, reverse-transcription and PCR analysis

Total RNA was extracted from snap frozen SH-SY5Y and/or T cell pellets using the innuPREP RNA Mini Kit from Analytic Jena AG, following the manufacturer’s recommendations. Five hundred nanograms of the isolated RNA was reverse transcribed using the M-MLV reverse transcriptase from Promega. Real-time PCR was performed using the Premix Ex Taq™ (probe qPCR, TaKaRa) in duplicates to determine the mRNA levels of the interferon (IFN)-stimulated genes (ISG) *IFIT1*, *IFI27*, *IFI44*, *IFI44L*, *ISG15*, *MX1*, *RSAD2* as well as *GAPDH* (housekeeping) using FAM-tagged TaqMan™ Gene Expression Assays purchased from Thermo Scientific following the manufacturer’s guidelines. The cycle threshold (Ct) values for target genes were converted to values of relative expression using the relative quantification (RQ) method (2-ΔΔCt). Target gene expression was calculated relative to Ct values for the GAPDH control housekeeping genes. In some experiments, interferon (IFN) scores were calculated as the median of the RQ of the seven ISG over a single calibrator control following the procedure of Rice et al. as in ^11^. Semi-quantitative PCR was conducted on SH-SY5Y cDNA to amplify overexpressed HA-*PSMC5* using a forward (5’-ATGGCGCTTGACGGACCAGAGCAGA-3’) primer binding to *PSMC5* and a reverse primer (5’-GACAGTGGGAGTGGCACCTTCCAGGGTCAAGG-3’) binding to the polyadenylation signal of the pcDNA3.1/Zeo(+) expression vector. Amplified cDNA products were resolved on 1.7% agarose gels, stained with 1.0 mg/ml GelRed®, and visualized by a UV transilluminator at 312 nm.

### Native-PAGE and in-gel activity assays

Resting T cells from control and NDD affected individuals with *PSMC5* variants were subjected to non-denaturing protein extraction by three cycles of freeze (liquid nitrogen) and thaw using TSDG buffer (10 mM Tris pH 7.0, 10 mM NaCl, 25 mM KCl, 1.1 mM MgCl_2_, 0.1 mM EDTA, 1 mM DTT, 1 mM ATP, 1 mM NaN_3_ and 20 % Glycerol). Samples were then centrifuged (14,000xg) for 15 minutes at 4°C and protein content in supernatants was determined using a standard Bradford assay. Twenty micrograms of whole-cell extract proteins were separated on 3-12% gradient Bis-Tris gels (Invitrogen) and in-gel activity assay was performed to visualize proteasome complexes by incubating gels with 0.1 mM fluorogenic peptide (LLVY-AMC, purchased from Bachem) for 30 minutes at 37°C. In addition, gels were blotted onto PVDF membranes (200V, 400 mA, 1h at 4°C under wet conditions) which were subsequently incubated with primary antibodies directed against the proteasome subunits PSMC5/Rpt6 (Enzo Life Sciences, clone p45-110) and α6 (Enzo Life Sciences, clone MCP20) overnight at 4°C. Antibody binding was then detected using an anti-mouse horseradish peroxidase (HRP)-labelled secondary antibody and chemiluminescence (Biorad).

### Analysis of gene expression through NanoString

A total of 100 nanograms of RNA was extracted from T cells sourced from both control and affected samples. The isolated RNA underwent hybridization using the NanoString nCounter® Human AutoImmune Profiling Panel. Following this, gene expression quantification adhered to the manufacturer’s instructions. The acquired data were subsequently normalized to housekeeping genes in accordance with the manufacturer’s guidelines.

### Lipid profile analysis

Lipid profiling was achieved by high-resolution mass spectrometry coupled to an reversed phase UHPLC system. Lipid extraction from donor-derived cells was performed according to ^17^ with small modifications. In brief, 225 μL of cold MeOH w/ 0.01 % BHT was added to an PBMC cell pellet, vortexed and mixed by pipetting up and down, and pulse sonicated using a tip sonicator for 30s on ice. After adding 3 μL of the EquiSPLASH (Avanti Polar Lipids, Alabaster/AL, USA) and 750 μL of cold MTBE the mixture was incubated for 1 h at 4 °C in a thermomixer at 650 rpm. After adding 188 μL of ultrapure water the sample was centrifuged at 10,000 g for 10 min. The upper (organic) layer (700 μL) was transferred to a vial and stored at 4 °C. The lower phase was extracted again with 400 µL MTBE and 10 µL acetic acid for 30 min at 4 °C (thermomixer), and 400 µL of the upper organic layer was transferred to first extract. After drying with sodium sulphate, the solvent was removed by nitrogen stream and stored at −80 °C until analysis.

After rehydration with chloroform/methanol/isopropanol (1:2:4; buffered with ammonium acetate pH 8), samples were analyzed in duplicate by a Vanquish UHPLC system equipped with an AccuCore C30 column (2.1×100 mm, 2.6 µm, Thermo Fisher Scientific) connected via nano-electrospray to an QExactive Plus high-resolution mass spectrometer in positive and negative mode (four injections per sample). Separation was achieved at 50 °C using a linear gradient built from: 60:40 water:acetonitrile (v/v, 10 mM ammonium formate/0.1 % formic acid, buffer A) and 90:10 Isopropanol:Acetonitrile (v/v, 10 mM ammonium formate/0.1 % formic acid, buffer B). Composition changed from 20 % B to 99 % B within 29 min. Mass spectra were recorded in data-dependent acquisition (DDA, loop count 15) mode with a resolving power of 70.000 in MS1 mode and 17.500 in MS2. Normalized collision energy for MS2 fragmentation was set to 25. Spray voltage on a HESI II ion source was set to 2.80 kV with adjusted spray parameters for stable spray conditions in each polarity. MS raw data were analyzed using LipidSearch (Version 4.1.16; Thermo Fisher Scientific). Lipid identification workflow was set to a precursor tolerance of max 5 ppm, a product tolerance of max 7 ppm and an m-score threshold of 2.0. The top rank filter was enabled as well as the main node filter was set to main isomer peak. Fatty acid priority was enabled. ID quality filter was set to A-B, allowing only identifications based on the whole structure including head group, glycerol backbone and fatty acid chains. All lipid classes were selected for identification. Ion adducts for positive mode measurements were: +H, +NH4, +2H and for negative mode measurements –H, +HCOO- and - 2H. After identification, lipid species were filtered for peak quality (over 0.8) and measured mass deviations (between - 1 and 1 ppm).

## Results

### Rare pathogenic *PSMC5* variants cause a syndromic neurodevelopmental disorder

We identified 38 unrelated individuals who exhibited similar clinical features and carried a rare single nucleotide variant (SNV) or indel in *PSMC5*. Thirty cases were *de novo*, five assumed *de novo*, two inherited from an affected mother, and one recessive. In total, we identified 23 distinct variants, four of them recurring (**Table 1**). Most variants were missense (18/23) and impacted regions highly conserved from human to yeast (**Fig. S1**). They were predominantly located in the “ATPases Associated with diverse cellular Activities (AAA-ATPase)” domain (14/23); the recurring variants NM_002805.6: c.959C>G p.(Pro320Arg) (7/38 subjects) and c.973C>T p.(Arg325Trp) (8/38) were located near the end of the domain (**Fig. 1A**).

**Table 1.** Main characteristics of the *PSMC5* variants identified in the affected individuals included in the study.

| Variant | cDNA change | Protein change | Inheritance | Variant databases | Mobidetails | Altered splicing | Subjects |
| --- | --- | --- | --- | --- | --- | --- | --- |
| V1 | c.166+3del | p.? | *de novo* | - | 212698 | + | S1 |
| V2 | c.206A>G | p.(Gln69Arg) | *de novo* | - | 212702 | - | S2 |
| V3 | c.414G>C | p.(Met138Ile) | *de novo* | - | 212704 | - | S3 |
| V4 | c.548C>T | p.(Pro183Leu) | assumed *de novo* | - | 212706 | - | S4 |
| V5 | c.587del | p.(Lys196Argfs*29) | maternally inherited* | - | 306340 | - | S5 |
| V6 | c.601C>T | p.(Arg201Trp) | assumed *de novo*/ *de novo* | - | 212708 | - | S6, S7 (2) |
| V7 | c.605C>T | p.(Ala202Val) | *de novo* | - | 212710 | - | S8 |
| V8 | c.620C>T | p.(Thr207Met) | recessive (homozygous) | gnomAD(1); ALFA(1);<br>rs749452987 | 212712 | - | S9 |
| V9 | c.647G>A | p.(Gly216Asp) | *de novo* | - | 212714 | - | S10 |
| V10 | c.662A>G | p.(Gln221Arg) | assumed *de novo* | - | 212716 | + | S11 |
| V11 | c.749A>T | p.(Glu250Val) | *de novo* | - | 212718 | - | S12 |
| V12 | c.749A>G | p.(Glu250Gly) | *de novo* | - | 212720 | - | S13 |
| V13 | c.754G>A | p.(Asp252Asn) | *de novo* | - | 212722 | - | S14, S15 (2) |
| V14 | c.772C>T | p.(Arg258Trp) | *de novo* | gnomAD(2); rs11543211 | 212724 | - | S16 |
| V15 | c.773G>A | p.(Arg258Gln) | maternally inherited* | rs781522827, but no MAF | 274699 | - | S17 |
| V16 | c.845G>A | p.(Gly282Asp) | *de novo* | - | 307153 | - | S18 |
| V17 | c.959C>G | p.(Pro320Arg) | *de novo* | - | 212726 | - | S19-S25 (7) |
| V18 | c.959C>A | p.(Pro320His) | assumed *de novo* | BRAVO (1) ; rs1349754006 | 212728 | - | S26 |
| V19 | c.968_969+1del | p.? | *de novo* | - | 307156 | + | S27 |
| V20 | c.970-2A>G | p.? | assumed *de novo* | - | 212730 | + | S28 |
| V21 | c.973C>T | p.(Arg325Trp) | *de novo* | - | 212732 | - | S29-S36 (8) |
| V22 | c.1103T>C | p.(Met368Thr) | *de novo* | gnomAD (1) | 212734 | - | S37 |
| V23 | c.1182_1183del | p.(Asp394Glu fs*2) | *de novo* | - | 212736 | - | S38 |

2 RefSeq transcript used for *PSMC5* is NM\_002805.6. \*inherited from an affected mother
3 Variant database used: gnomAD v2.1.1/v3.1.2, dbSNP v154, Clinvar, ALFA v20201027095038, BRAVO/TOPmed data freeze 8; MAF: minor allele frequency; 4 tools used for assessing effect on splicing: SPIP, SpliceSiteFinder, MaxEntScan, NNSplice, GeneSplicer; Intervar: Pat= pathogenic
5 The access to detailed predictions for variant XXXXXX (e.g. 212698) using Mobidetails is as follows: https://mobidetails.iurc.montp.inserm.fr/MD/variant/XXXXXX

**Table 2.** Clinical features of the subjects with PSMC5 variants and indels (1/2).

| Subject | S1 | S2 | S3 | S4 | S5 | S6 | S7 | S8 | S9 | S10 | S11 | S12 | S13 | S14 | S15 | S16 | S17 | S18 | S19 | S20 |
| --- | --- | --- | --- | --- | --- | --- | --- | --- | --- | --- | --- | --- | --- | --- | --- | --- | --- | --- | --- | --- |
| Variant | V1 | V2 | V3 | V4 | V5 | V6 | V6 | V7 | V8 | V9 | V10 | V11 | V12 | V13 | V13 | V14 | V15 | V16 | V17 | V17 |
| Sexr | M | M | M | M | F | F | F | F | M | M | F | F | F | M | F | M | M | M | M | F |
| Age (in years) | 0-5 | 0-5 | 0-5 | 0-5 | 0-5 | 6-10 | 0-5 | 16-20 | 11-15 | 0-5 | 11-15 | 6-10 | 16-20 | 11-15 | 0-5 | 6-10 | 26-30 | 11-15 | 0-5 | 0-5 |
| Neurodevelopmental abnormality | + | + | + | + | + | + | + | + | + | + | + | + | + | + | + | + | + | + | + | + |
| Speech impairment (delay, absence) | + | + | + | - | + | + | + | + | ND | + | + | + | + | + | + | - | + | + | + | + |
| Abnormal facial shape | + | + | + | - | + | - | + | + | + | + | - | - | + | + | + | ND | - | - | + | + |
| Abnormal muscle tone | + | - | - | - | + | + | + | + | - | + | - | - | + | + | + | ND | - | - | + | + |
| Motor delay/impairment | + | - | + | + | + | + | + | + | + | + | - | + | + | - | + | ND | - | - | + | + |
| Ophthalmological findings | + | - | - | + | - | + | - | + | + | + | - | + | + | + | + | ND | ND | - | + | - |
| Intellectual disability | - | ND | ND | + | + | + | ND | - | - | ND | + | + | + | + | + | - | - | + | ND | ND |
| Abnormal behavior | - | - | + | + | + | + | - | + | + | - | + | - | - | + | - | + | + | + | + | - |
| Brain MRI abnormalities | - | ND | - | ND | ND | - | - | + | ND | + | ND | ND | + | + | - | ND | - | - | + | ND |
| Abnormal ear morphology | - | + | + | - | + | - | + | ND | + | + | - | - | + | + | - | ND | - | - | + | - |
| Other findings | - | - | + | + | + | + | + | + | - | - | + | - | - | + | + | + | + | - | - | - |
| Skeletal abnormalities (not involving skull) | - | - | + | - | + | - | + | + | + | - | - | - | + | - | + | - | - | - | - | - |
| Feeding difficulties | - | + | - | + | + | + | + | - | - | + | - | + | - | + | - | - | - | - | + | + |
| Abnormal weight | + | + | - | + | + | + | + | - | + | - | + | - | + | + | + | - | - | - | - | + |
| Abnormal length | + | + | + | + | + | - | + | - | + | - | + | - | + | - | - | - | - | - | - | + |
| Abnormal OFC | + | - | - | + | + | - | + | + | - | - | + | - | + | - | - | - | - | - | - | + |
| Other malformations | - | - | - | - | + | + | - | - | + | - | - | - | + | + | - | ND | - | + | + | - |
| Retrognathia/microretrognathia | - | + | + | - | - | - | - | ND | - | + | - | - | + | + | + | ND | - | - | - | - |
| Cardiac abnormalities | - | + | - | - | - | - | - | - | - | - | - | + | + | - | - | - | + | - | + | - |
| Seizures | + | - | - | - | - | + | - | - | - | - | - | - | + | - | + | - | + | + | - | - |
| Hearing loss | - | - | - | - | - | - | - | - | + | + | - | - | ND | - | + | - | - | - | - | - |
| Renal abnormalities | - | - | - | - | - | + | - | - | - | - | - | - | - | - | + | - | - | - | - | - |
| Genital abnormalities | - | - | + | + | - | - | - | - | - | - | - | - | - | - | - | - | - | - | - | + |
| Developmental regression | - | - | - | - | - | - | - | - | - | - | - | - | - | - | - | - | - | + | + | - |

**Table 2.**
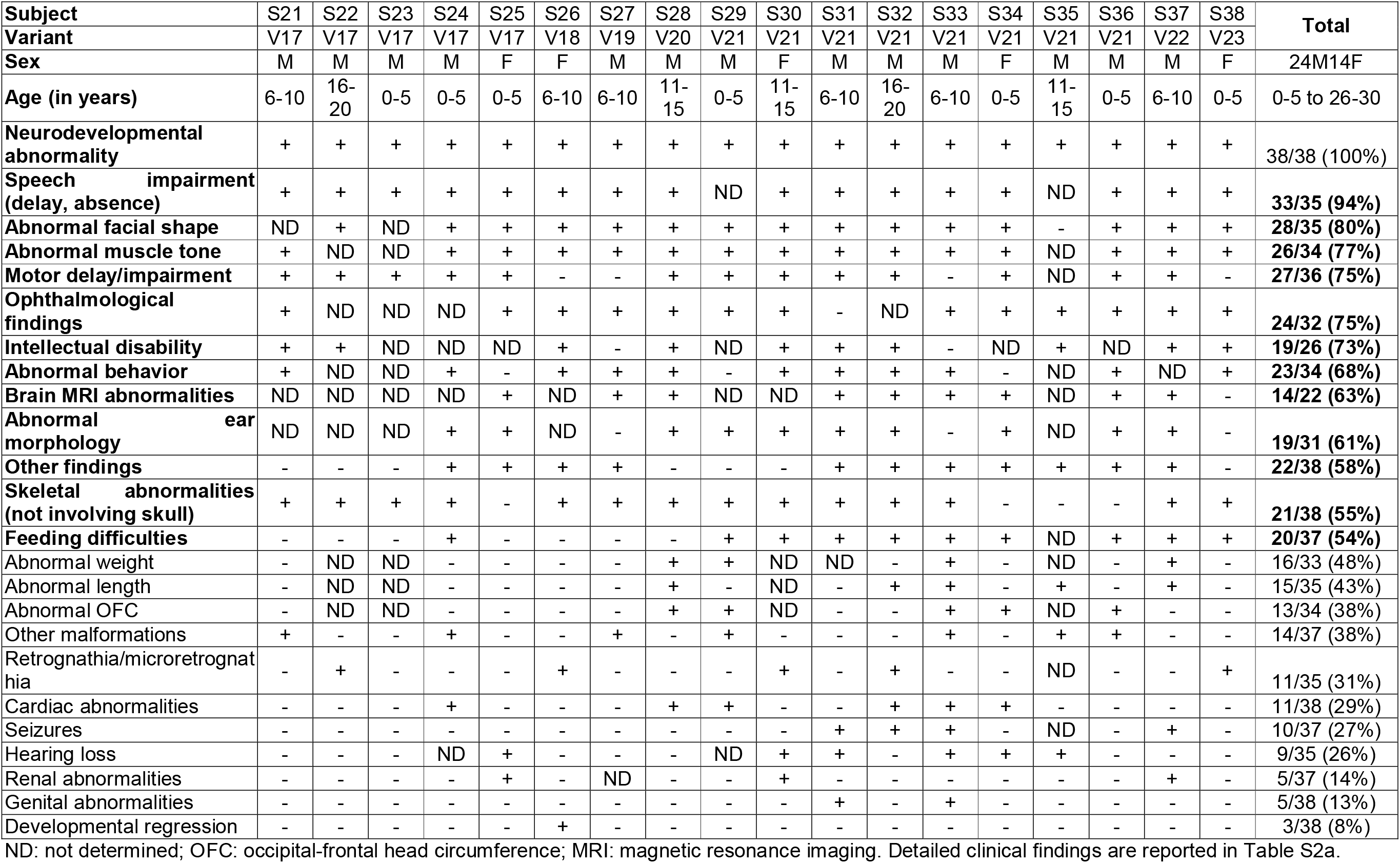
Clinical features of the subjects with PSMC5 variants and indels (2/2).

| Subject | S21 | S22 | S23 | S24 | S25 | S26 | S27 | S28 | S29 | S30 | S31 | S32 | S33 | S34 | S35 | S36 | S37 | S38 | Total |
| --- | --- | --- | --- | --- | --- | --- | --- | --- | --- | --- | --- | --- | --- | --- | --- | --- | --- | --- | --- |
| Variant | V17 | V17 | V17 | V17 | V17 | V18 | V19 | V20 | V21 | V21 | V21 | V21 | V21 | V21 | V21 | V21 | V22 | V23 |  |
| Sex | M | M | M | M | F | F | M | M | M | F | M | M | M | F | M | M | M | F | 24M14F |
| Age (in years) | 6-10 | 16-20 | 0-5 | 0-5 | 0-5 | 6-10 | 6-10 | 11-15 | 0-5 | 11-15 | 6-10 | 16-20 | 6-10 | 0-5 | 11-15 | 0-5 | 6-10 | 0-5 | 0-5 to 26-30 |
| Neurodevelopmental abnormality | + | + | + | + | + | + | + | + | + | + | + | + | + | + | + | + | + | + | 38/38 (100%) |
| Speech impairment (delay, absence) | + | + | + | + | + | + | + | + | ND | + | + | + | + | + | ND | + | + | + | 33/35 (94%) |
| Abnormal facial shape | ND | + | ND | + | + | + | + | + | + | + | + | + | + | + | - | + | + | + | 28/35 (80%) |
| Abnormal muscle tone | + | ND | ND | + | + | + | + | + | + | + | + | + | + | + | ND | + | + | + | 26/34 (77%) |
| Motor delay/impairment | + | + | + | + | + | - | - | + | + | + | + | + | - | + | ND | + | + | - | 27/36 (75%) |
| Ophthalmological findings | + | ND | ND | ND | + | + | + | + | + | + | - | ND | + | + | + | + | + | + | 24/32 (75%) |
| Intellectual disability | + | + | ND | ND | ND | + | - | + | ND | + | + | + | - | ND | + | ND | + | + | 19/26 (73%) |
| Abnormal behavior | + | ND | ND | + | - | + | + | + | - | + | + | + | + | - | ND | + | ND | + | 23/34 (68%) |
| Brain MRI abnormalities | ND | ND | ND | ND | + | ND | + | + | ND | ND | + | + | + | + | ND | + | + | - | 14/22 (63%) |
| Abnormal ear morphology | ND | ND | ND | + | + | ND | - | + | + | + | + | + | - | + | ND | + | + | - | 19/31 (61%) |
| Other findings | - | - | - | + | + | + | + | - | - | - | + | + | + | + | + | + | + | - | 22/38 (58%) |
| Skeletal abnormalities (not involving skull) | + | + | + | + | - | + | + | + | + | + | + | + | + | - | - | - | + | + | 21/38 (55%) |
| Feeding difficulties | - | - | - | + | - | - | - | - | + | + | + | + | + | + | ND | + | + | + | 20/37 (54%) |
| Abnormal weight | - | ND | ND | - | - | - | - | + | + | ND | ND | - | + | - | ND | - | + | - | 16/33 (48%) |
| Abnormal length | - | ND | ND | - | - | - | - | + | - | ND | - | + | + | - | + | - | + | - | 15/35 (43%) |
| Abnormal OFC | - | ND | ND | - | - | - | - | + | + | ND | - | - | + | + | ND | + | - | - | 13/34 (38%) |
| Other malformations | + | - | - | + | - | - | + | - | + | - | - | - | + | - | + | + | - | - | 14/37 (38%) |
| Retrognathia/microretrognathia | - | + | - | - | - | + | - | - | - | + | - | + | - | - | ND | - | - | + | 11/35 (31%) |
| Cardiac abnormalities | - | - | - | + | - | - | - | + | + | - | - | + | + | + | - | - | - | - | 11/38 (29%) |
| Seizures | - | - | - | - | - | - | - | - | - | - | + | + | + | - | ND | - | + | - | 10/37 (27%) |
| Hearing loss | - | - | - | ND | + | - | - | - | ND | + | + | - | + | + | + | - | - | - | 9/35 (26%) |
| Renal abnormalities | - | - | - | - | + | - | ND | - | - | + | - | - | - | - | - | - | + | - | 5/37 (14%) |
| Genital abnormalities | - | - | - | - | - | - | - | - | - | - | + | - | + | - | - | - | - | - | 5/38 (13%) |
| Developmental regression | - | - | - | - | - | + | - | - | - | - | - | - | - | - | - | - | - | - | 3/38 (8%) |

ND: not determined; OFC: occipital-frontal head circumference; MRI: magnetic resonance imaging. Detailed clinical findings are reported in Table S2a.

Our three-dimensional structural analysis showed notably that 13 of the variants primarily located in the lower ATPase domain interface with neighboring base ATPase subunits PSMC1/Rpt2 and PSMC4/Rpt3, and the substrate in its engaged state (**Figs. 1,S2**). They are likely to affect proteasome function in multiple ways, by disrupting proteasomal complex formation, impairing the transition from substrate-free to substrate-engaged states of the 26S proteasome, affecting ATP binding and hydrolyzation, hindering substrate processing, or perturbing protein folding and subunit incorporation (**Figs. 1,S2**). Overall, these findings, detailed in supplemental notes, align with the criteria that would classify of all 23 variants as pathogenic according to the American College of Medical Genetics and Genomics (ACMG) guidelines if *PSMC5* was an OMIM morbid gene (**Table S1**).

Abnormal neurodevelopment was evident in all subjects, with developmental delay in 35/38 (92%) cases (**Tables 2-S2b**). Neurological manifestations included speech absence or delay (33/35; 94%), abnormal muscle tone (26/34; 76%), motor delay or impairment (27/36; 75%), intellectual disability (19/26 (73%)), abnormal behavior (23/34; 68%, including autism spectrum disorder (ASD) and attention deficit hyperactivity disorder (ADHD)), and seizures (10/37; 27%) (**Tables 2-S2**). Brain magnetic resonance imaging revealed frequent but varied abnormalities in the examined subjects (14/22; 64%). The phenotype appeared syndromic (**Fig. 2A; Table S2b/S2c**), with notable non-neurological findings such as ophthalmological anomalies (24/32; 75%), skeletal malformations (21/38; 55%), feeding difficulties (20/37; 54%), cardiac abnormalities (11/38; 29%), hearing loss (9/35; 26%, conductive in four individuals, sensorineural and mixed in one patient each), kidney abnormalities (5/37; 14%) and genital abnormalities (5/38; 13%).

### Subjects with *PSMC5* variants present a similar facial gestalt

Most affected individuals exhibited abnormal facial shape (28/35; 80%), often accompanied by craniofacial abnormalities, including microcephaly (12/32; 38%) or abnormality of the mandible (11/35; 31%, including micrognathia, retrognathia and prognathia) (**Fig. 2B**). However, the manifestations were heterogeneous and did not allow conclusive diagnosis of the disorder through strandard clinical assessment. Nonetheless, facial analysis with GestaltMatcher^15^ suggested a recognizable facial gestalt among *PSMC5* subjects. A strong facial resemblance was indeed observed between the 13 *PSMC5* subjects tested, 95% of subject combinations having mean pairwise distances below the threshold (*c=0.915*) (**Fig. S4**). Besides, the analysis by pairwise comparison matrix extended to 7,459 images from 449 disorders in GMDB revealed that 10 of 13 *PSMC5* subjects (S3/9/10/12/13/19/30-33) matched at least another *PSMC5* subject with a rank below 50 (**Fig. 2C**). Subject pairs/trios (S9 and S33), (S10, S19 and S31), and (S13 and S30) were highly similar, with ranks below five, indicating again strong resemblance..

### Knockdown (KD) of *Psmc5* alters fly reversal learning and excitatory/inhibitory balance of rat hippocampal neurons

Pan-neuronal KD of *Drosophila melanogaster* highly conserved orthologue of *PSMC5*, *Rpt6*, had no significant effect on classical olfactory learning where flies learn an association between odor and foot shock (Tukey, p=0.3059; N=4; **Fig. 3A**). However, they presented with significant defect in the ability to learn novel association following initial training (a different behavior paradigm known as reversal learning-see method for details) (Tukey, p<0.0001; N=4; **Fig. 3B**). Importantly, this is similar to what we previously reported with another proteasome subunit gene, *PSMC3*^11^.

**Figure 3:**
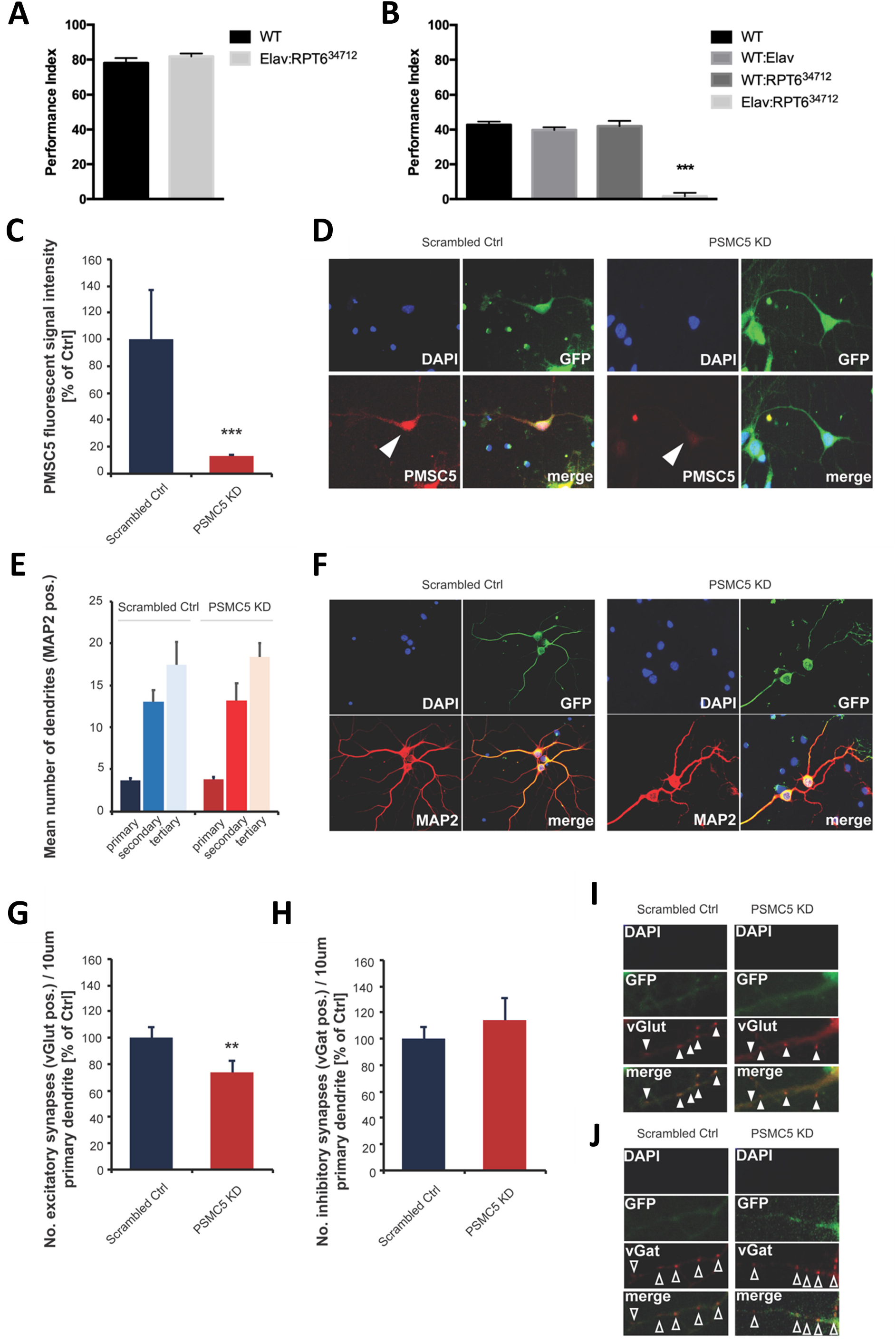
Suppression of *Psmc5* leads to changes in reversal learning in flies and disrupts the balance between excitatory and inhibitory signals in rat hippocampal neurons. *A.* Pan-neuronal knockdown of Rpt6 in *Drosophila* did not significantly affect normal olfactory learning (p=0.3059, n=4). *B.* When faced with the more difficult task of reversal olfactory learning, knocking down Rpt6 pan-neuronally in *Drosophila* resulted in a significant decrease in performance (p<0.0001, n=4). *(C-J)* Primary rat hippocampal neurons infected DIV1-DIV14 with scrambled shRNA or PMSC5 knockdown (KD) shRNA delivering lenti-particles that co-express GFP. *C.* Assessment of PMSC5 fluorescent signal intensity in neurons at DIV14 shows a significant reduction in cells infected (GFP positive) with *PMSC5* knockdown (KD) compared to cells infected with scrambled shRNA (n=10, Mann Whitney test, p = 0.0002). *D.* Exemplary images of neurons infected with scrambled shRNA (left) or PMSC5 shRNA (right) lenti-particles. PMSC5 labeling (arrow) is significantly reduced, confirming the KD of PMSC5. *E-F.* Infected neurons stained for MAP2 to visualize neuronal morphology. *E.* No significant difference in dendritic branching measured by the mean number of primary, secondary, and tertiary dendrites per neuron at DIV14 was found comparing scrambled Ctrl and PMSC5 KD conditions (n=6). *F.* Exemplary images of neurons infected with scrambled shRNA (left) or PMSC5 shRNA (right) lenti-particles. MAP2 labeling is shown in red. *G-J.* Infected neurons stained for vGlut to visualize excitatory presynaptic compartments or vGat to visualize inhibitory presynapses. *G.* The number of vGlut positive puncta along the primary dendrites was measured at DIV14. In *Psmc5* KD conditions, a significant reduction was observed compared to scrambled Ctrls (n=5, Mann Whitney test, p = 0.0045). *H.* The number of vGat positive puncta along the primary dendrites was not significantly altered (n=5, Mann Whitney test, p = 0.2988). *I-J.* Representative images of primary dendrites of neurons infected with scrambled shRNA (left) or *Psmc5* shRNA (right) lenti-particles. VGlut (full arrow) positive signals per dendrite length are significantly reduced, while VGat (open arrow) positive signals per dendrite length are not altered after *Psmc5* KD.

We studied the effects of *Psmc5* loss on primary rat hippocampal neurons by gene KD, using *Psmc5*-specific shRNA or scrambled shRNA for controls (scrambled Ctrl) along with coexpressed GFP delivered via lenti-virus infection of cells at day 1 of development (**Fig. 3C-J**). After 14 days in culture, a significant reduction of PSMC5 levels was measured by fluorescent labelling with PSMC5-specific antibodies (**Fig. 3C-D**). Notably, under control conditions, PSMC5 labelling appeared predominantly in neurons and much less in glial cells.

To assess the general morphology of neurons developing under *PMSC5* KD, we measured their dendritic arborization at days *in vitro* (DIV) 14. No significant differences were observed in *PSMC5* KD neurons compared to controls (**Fig. 3E-F**). We also measured synapse density of excitatory synapses (visualized using vGlut as excitatory presynaptic marker) and inhibitory synapses (visualized using vGat as inhibitory presynaptic marker) (**Fig. 3G-J**). After 14 days, *PSMC5* loss led to a significantly reduced number of vGlut positive puncta along primary dendrites (**Fig. 3G-I**), whereas the number of vGat positive puncta remained unchanged. (**Fig. 3H-J**). This indicates an excitation/inhibition (E/I) imbalance due to PSMC5 loss in neurons.

### Overexpression of *PSMC5* affects neuronal morphology in mouse

A subset of identified missense variants was assessed for their impact on neuronal morphology through overexpression in primary mouse hippocampal neurons. Neurite length of mouse hippocampal cells was significantly increased during the neurodevelopmental phase (DIV3) by overexpression of protein product PSMC5-WT compared to empty vector control (**Fig. 4A-B**), suggesting a potential role of *Psmc5* in promoting neurite outgrowth. Overexpression of variants p.(Arg201Trp), p.(Pro320His) and p.(Arg325Trp) yielded the same effect as PSMC5-WT, suggesting that in this assay these variants do not alter the effect of overexpression of *PSMC5*. By contrast, overexpression of variants p.(Ala202Val), p.(Thr207Met), p.(Pro320Arg) and PSMC5-Δe×10 (c.970-2A>G) showed reduced neurite length compared to PSMC5-WT (**Fig. 4A-B**); this effect, similar to the one induced by the empty vector, suggest that these variants have a loss of function (LoF) effect in the assay. Notably, no effect on arborization was induced by PSMC5-WT and none of the variants had impact on arborization compared to the empty vector control (**Fig. 4B**).

**Figure 4:**
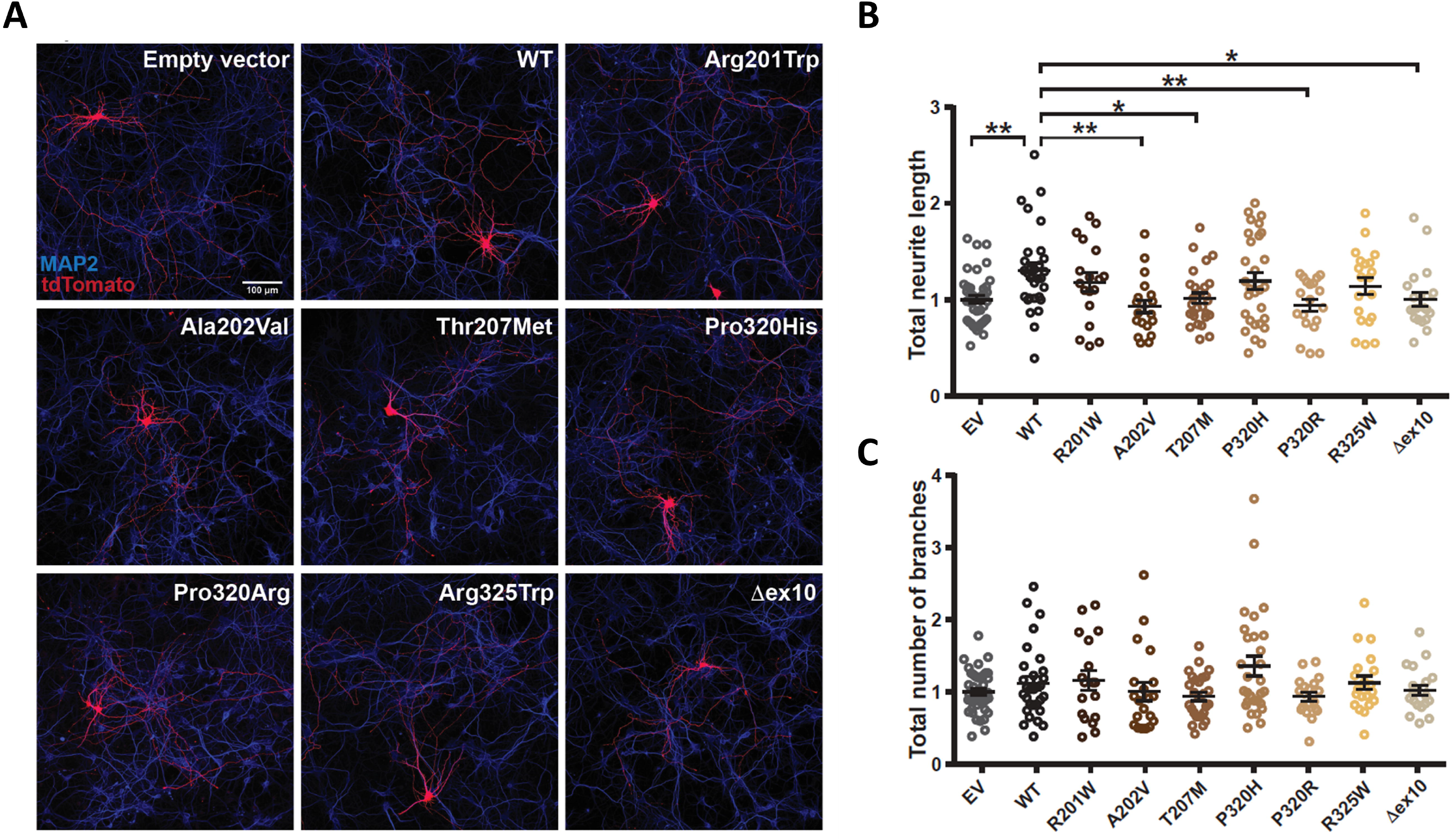
Overexpression of *PSMC5* affects neuronal morphology. ***A.*** Representative images of primary hippocampal neurons transfected with the empty vector control (EV), *PSMC5*-WT or the different *PSMC5* variants. Transfected cells are marked in red using tdTomato, while the neuronal marker MAP2 is stained in blue. ***B.*** Analysis of the total neurite length in the different conditions reveal: (i) a significant increase induced by overexpression of PSMC5-WT compared to empty vector control (One-way ANOVA, F=3.296, p= 0.0014; empty vector versus PSMC5-WT, p=0.0046, Dunnet’s multiple comparison test); (ii) a similar effect as PSMC5-WT induced by overexpression of variants p.(Arg201Trp), p.(Pro320His) and p.(Arg325Trp) (PSMC5-WT versus p.(Arg201Trp), p=0.8; PSMC5-WT versus p.(Pro320His), p=0.8; PSMC5-WT versus p.(Arg325Trp), p=0.5, Dunnet’s multiple comparison test); (iii) a decrease induced by overexpression of variants p.(Ala202Val), p.(Thr207Met), p.(Pro320Arg) and PSMC5-Δe×10, compared to PSMC5-WT (PSMC5-WT versus p.(Ala202Val), p=0.0031; PSMC5-WT versus p.(Thr207Met), p=0.02; PSMC5-WT versus p.(Pro320Arg), p=0.006; PSMC5-WT versus PSMC5-Δe×10, p=0.03, Dunnet’s multiple comparison test). The effects of these variants were indistinguishable from the empty vector control (empty vector versus p.(Ala202Val), p=0.4; empty vector versus p.(Thr207Met), p=0.9; empty vector versus p.(Pro320Arg), p=0.9; empty vector versus PSMC5-Δe×10, p=0.9, Dunnet’s multiple comparison test). ***C.*** Analysis of the arborization showed no differences between conditions compared to the empty vector control (One-way ANOVA, F=2.179, p= 0.03; empty vector versus PSMC5-WT, p=0.9, PSMC5-WT versus p.(Arg201Trp), p=0.9; PSMC5-WT versus p.(Ala202Val), p=0.9; PSMC5-WT versus p.(Thr207Met), p=0.6; PSMC5-WT versus p.(Pro320His), p=0.3; PSMC5-WT versus p.(Pro320Arg), p=0.9; PSMC5-WT versus p.(Arg 325Trp), p=0.9; PSMC5-WT versus PSMC5-Δe×10, p=0.9, Dunnet’s multiple comparison test). Error bars indicate SEM, n (number of neurons traced): EV=40, PSMC5-WT=30, p.(Arg201Trp)=18 p.(Ala202Val)=20, p.(Thr207Met)=27, p.(Pro320Arg)=29, p.(Pro320Arg)=19, p.(Arg325Trp)=20 and PSMC5-Δe×10=20. *p<0.05; **p<0.01. Scale bar: 100μm.

### The *PSMC5* variants associated with NDD do not equally impact the PSMC5/Rpt6 subunit steady-state expression and subsequent incorporation into 26S proteasomes

To analyze the ability of PSMC5/Rpt6 variant subunits to integrate 26S proteasome complexes, we ectopically expressed 13/23 identified alterations as N-terminally HA tagged-*PSMC5* versions in SHSY-5Y neuroblastoma cells, as previously described^11^.

While the plasmid-driven production of *PSMC5* transcripts was similar between the wild-type and the 13 variant cell lines (**Fig. S5A**), steady-state protein expression levels of PSMC5/Rpt6 were differentially impacted by the variants: they were reduced profoundly by p.(Ala202Val), moderately by p.(Pro183Leu), p.(Arg201Trp), p.(Gly216Asp), p.(Glu250Val) and p.(Pro320Arg), and mildly by p.(Arg258Trp), p.(Pro320His) and p.(Arg325Trp), whereas they were unaltered by p.(Thr207Met), p.(Met368Thr) and p.(Asp394Glufs*2) (**Fig. S5B**). Overall, analyses of the 13 *PSMC5* variants examined revealed no significant impact on *in vitro* PSMC5/Rpt6 abundance when compared to control (**Fig. S5B**, right panel), indicating that haploinsufficiency is unlikely the main driver of variant pathogenicity.

We also observed distinct effects of the 13 *PSMC5* variants on proteasome assembly in this *in vitro* assay. Notably, p.(Ala202Val) prevented PSMC5 incorporation into 19S, 26S, and 30S complexes (**Fig. S5C**), due to the instability of the HA-PSMC5/Rpt6 full-length protein (**Fig. S5B**). Conversely, despite their stable expression in SH-SY5Y cells (**Fig. S5B**), p.(Met368Thr) and p.(Asp394Glufs*2) accumulated in 19S precursors and fully assembled 19S particles without associating with 26S and/or 30S proteasome complexes (**Fig. S5C**). Unlike variants p.(Gly216Asp), p.(Gln221Arg), p.(Arg201Val) and p.(Arg258Trp) which showed unchanged incorporation efficiency compared to their wild-type counterpart, p.(Pro183Leu), p.(Glu250Val), p.(Pro320His) and p.(Pro320Arg) only minimally assembled into mature proteasomes, (**Fig. S5C**). Surprisingly, p.(Thr210Met) had a higher propensity to integrate 19S-capped proteasomes than the wild-type. Densitometric analysis showed that the 13 *PSMC5* variants studied had similar incorporation into 19S, 26S, and/or 30S complexes compared to their wild-type counterparts (**Fig. S5C**, lower panel). These findings underscore the varied effects of these variants on subunit expression, protein level stability, and incorporation into mature 26S/30S proteasome complexes.

### Most *PSMC5* variants do not lead to haploinsufficiency but result in severe proteasome assembly defects

In T cells from affected subjects, we observed that,. except p.(Pro320Arg), no *PSMC5* variants reduced PSMC5/Rpt6 levels compared to control samples (**Fig. 5A**). The unchanged levels of PSMC5/Rpt6 in T cells from S12 [p. (Glu250Val)] and S32 [(p. Arg325Trp)], were discordant with *in vitro* results that had hinted at variant-induced protein instability in SHSY-5Y cells (**Fig. S5B**). This observation suggests a full compensation by the wild-type allele and counters haploinsufficiency as a cause of the disorder. Notably, while the PSMC5/Rpt6 steady-state protein expression levels were comparable between control and subjects, all T cell specimens with *PSMC5* variants showed extra lower-migrating PSMC5/Rpt6 species compared to controls (**Fig. 5A, lower panel**), possibly due to a dysregulation of *PSMC5* transcript splicing.

**Figure 5:**
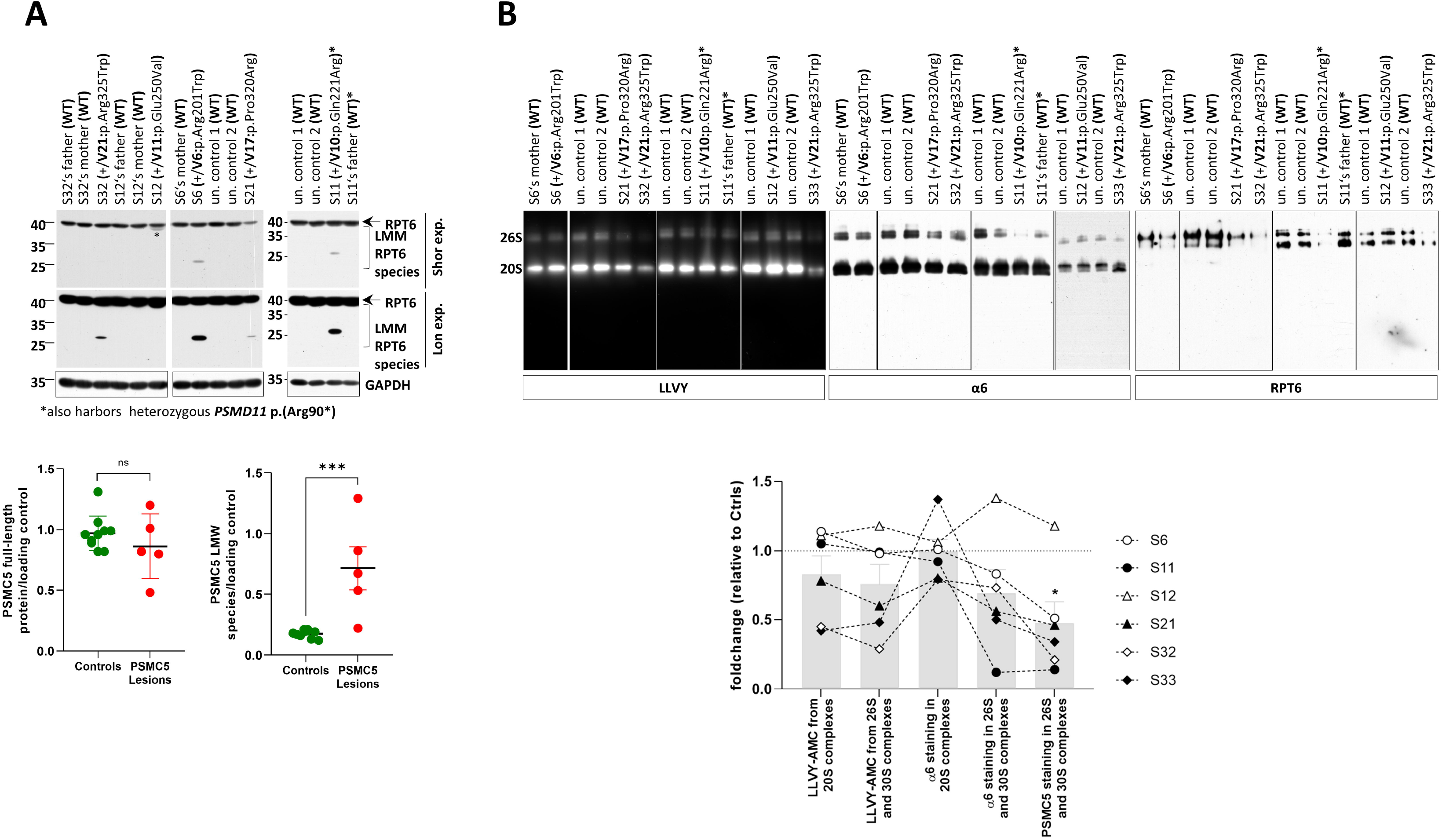
T cells from NDD subjects carrying *PSMC5* heterozygous variants exhibit abnormalities in their proteasome expression and/or activity profiles. *A*. T cells expanded from PBMC isolated from NDD subjects S6/11/12/21/32 as well as related controls (father and/or mother) and heathy donors (noted ‘un. control’ for unrelated controls) were lysed in RIPA buffer prior to SDS-PAGE/western-blotting analysis using antibodies specific PSMC5/RPT6 and GAPDH (loading control). Arrow points to the expected size of full-length endogenous PSMC5/RPT6 detected in all samples, at approximately 45 kDa. PSMC5/Rpt6 staining of T cell lysates from subjects 6/11/21/32 demonstrates additional shorter PSMC5/Rpt6 immunoreactive band migrating at ∼30kDa (denoted as a low-molecular mass (LMM) species by the square brackets) following prolonged exposure times, whereas T cells from subject S12 exhibited an extra PSMC5/Rpt6 signal running a little bit lighter than PSMC5/Rpt6 expected size just below 40 kDa (indicated by an asterisk). Lower panel: the RPT6 and GAPDH immunoreactive bands were quantified by densitometry. Data are presented as RPT6 full-length and low molecular weight (LMW) species on GAPDH ratio mean values ± SD for the control (n=9) and patients (PSMC5 lesions, n=5) groups, as indicated. Statistical analysis was performed using the Mann-Whitney U test where *** indicates *p*<0.001. *B*. Resting T cells from NDD subjects S6/11/12/21/32/33 as well as related (subject’s father and/or mother, as indicated) and unrelated (healthy donor) control T cells were analyzed for proteasome function and abundance. They were subjected to non-denaturing protein extraction using TSDG buffer prior to native-PAGE/western-blotting analysis using monoclonal antibodies specific for the α6 and PSMC5/RPT6 subunits, as indicated. Proteasome complexes (20S and 26S) were also visualized by in-gel activity assay using the Suc-LLVY-AMC fluorogenic peptide reflecting chymotrypsin-like activity. Lower panel: the LLVY-AMC fluorescent signals as well as the α6 and RPT6 immunoreactive bands in 30S and/or 26S proteasome complexes were quantified by densitometry and presented as fold change values in patients S6, S11, S12, S21, S32 and S33 versus the control group for each gel whose densitometry measurements were set to 1 (grid line). Columns indicate foldchange mean values ± SD of the patient group (n=6) for LLVY-AMC, α6 and RPT6 in 30S and/or 26S complexes, as indicated. Statistical analysis was performed using the Mann-Whitney U test where * indicates p<0.05.

In line with *in vitro* findings, the T cells of subjects S6 [p.(Arg201Trp)], S21 [p.(Pro320Arg)], S32 [p.(Arg325Trp)], and S33 [p.(Arg325Trp)] exhibited reduced incorporation of PSMC5/Rpt6 into 26S and 30S proteasome complexes, as evidenced by reduced staining intensity of PSMC5/Rpt6 in the 19S-capped proteasomes (**Fig. 5B**). However, for two other *PSMC5* variants, there was a discrepancy between *in vivo* and *in vitro* results: (i) p.(Gln221Arg) was associated with decreased PSMC5/Rpt6 subunit incorporation in T cells from S11 (**Fig. 5B**), potentially influenced by a second proteasomal variant, NM_002815.3:c.268C>T p.(Arg90*) in *PSMD11*, inherited from a moderately affected father. Although this additional variant was not considered in our *in vitro* assays, its impact on 26S proteasome assembly seems evident, as demonstrated by the reduced amounts of 30S proteasomal complexes and the decreased α6 staining intensity in T cells of S11’s father; (ii) conversely, p.(Glu250Val) did not impair PSMC5/Rpt6 assembly into 26S/30S proteasomal complexes in S12’s T cells (**Fig. 5B**), in contrast to the significant disruption observed in SHSY-5Y cells (**Fig. S5C**). In summary, *PSMC5* variants exhibit diverse behavior, with some of them failing to maintain stable incorporation into mature proteasomes.

### *PSMC5* variants profoundly disrupt protein homeostasis and lipid metabolism, and activate mitophagy in T cells from NDD subjects

To determine the effect of *PSMC5* variants on protein homeostasis, we evaluated the propensity of T cells from individuals S1/6/11/12/21/32 to form protein aggregates. We observed increased an aggresome formation in all subject samples (**Fig. 6A**), along with the accumulation of high molecular weight (HMW) ubiquitin-modified proteins in six samples (**Fig. 6B**). These findings demonstrate the loss-of-function nature of *PSMC5* variants, which prevent T cells from coping with proteotoxic stress.

**Figure 6:**
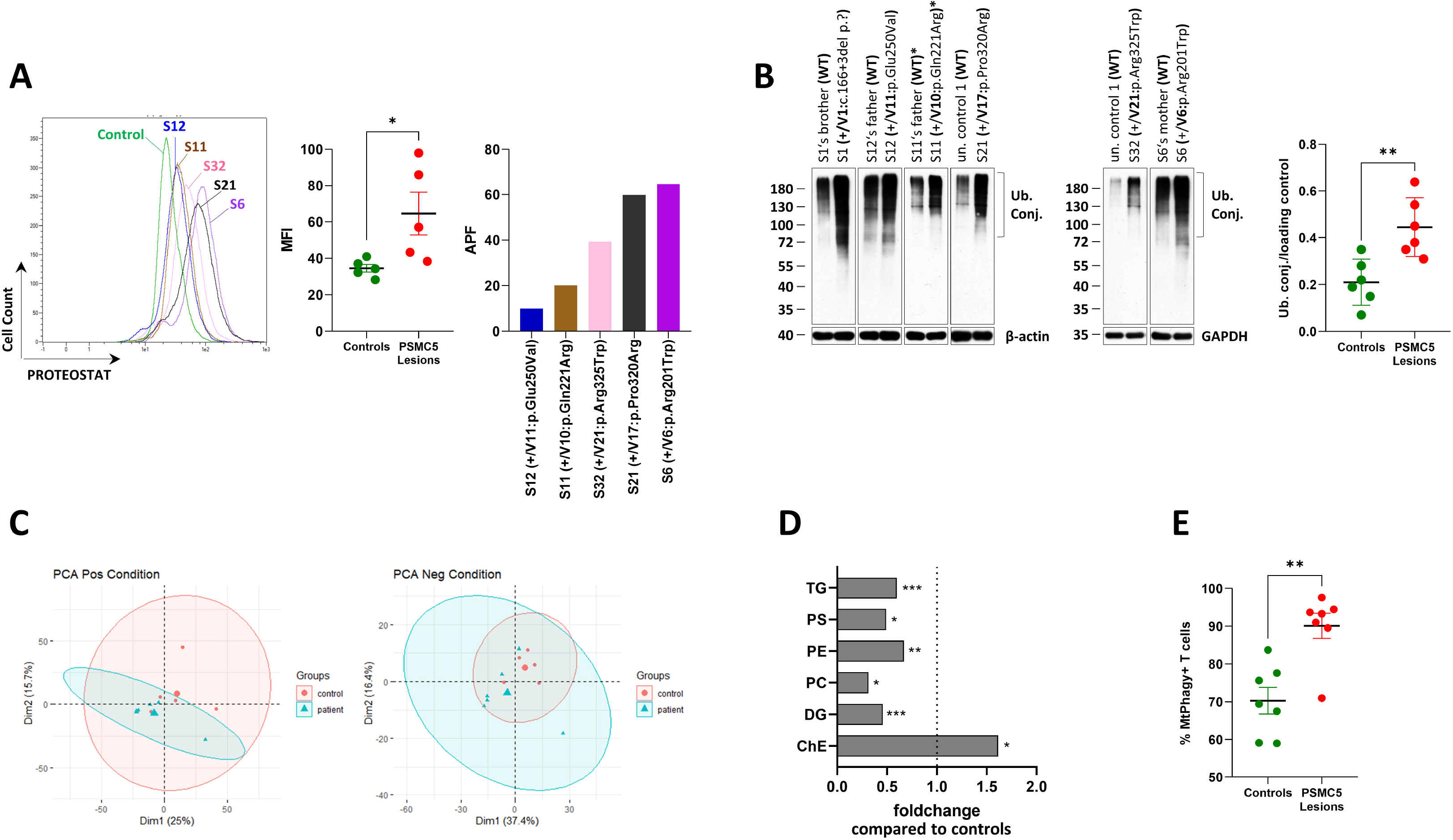
T cells from NDD subjects with *PSMC5* heterozygous variants are characterized by disrupted protein homeostasis, increased mitophagy rates and a specific lipid signature. *A*. T cells expanded from PBMC isolated from NDD subjects S1/6/11/12/21/32 were stained with 1 µM of the PROTEOSTAT® dye prior to flow cytometry analysis using the B3 (PerCP-Vio 700) channel. PROTEOSTAT®, a dye that specifically intercalates into the cross-beta spine of quaternary protein structures typically found in misfolded and aggregated proteins. All T cells with PSMC5 variants exhibited increased aggresome formation, as evidenced by increased PROTEOSTAT® fluorescence intensity detected using flow cytometry. Shown is a representative histogram overlay plot of the six T cell patient samples over a control one (left panel) and the individual mean fluorescence intensity (MFI) values of the patient (*PSMC5* lesions, n=6) and control (n=6) groups (right panel) (\**p*<0.05, Mann-Whitney U test). *B*. T cells from NDD subjects S1/6/11/12/21/32, relatives (subject’s father, mother and/or brother) and unrelated controls (healthy donors) were subjected to RIPA-mediated protein extraction and subsequent SDS-PAGE/western-blotting using antibodies directed against ubiquitin, β-actin (loading control) and GAPDH (loading control), as indicated. Right panel: the ubiquitin, β-actin immunoreactive bands were quantified by densitometry and data are presented as ubiquitin/loading control mean values ±SD for both control (n=6) and patient (PSMC5 lesion, n=6) groups, as indicated. Statistical analysis was performed using the Mann-Whitney test with ** indicating p<0.01. *C.* In an attempt to deepen our understanding of the involvement of proteasomes in the regulation of lipid metabolism, we undertook an untargeted lipidomic analysis of T cells from seven unrelated *PSMC5* Subjects (S1, S6, S11, S12, S19, S21 and S33) together with their six relative controls (probands’ brother, father and/or mother). Principal component analysis (PCA) plots were first generated to visualize lipid distribution and identify specific patterns across control and patient samples with positive (left panel) and negative (right panel) ion modes. The control and patient groups are presented in red and blue, respectively. *D*. Fold change analysis showing the lipid classes undergoing significant changes in T cells derived from NDD patients when compared to healthy donors. *E*. T cells expanded from PBMC isolated from the six NDD subjects S1/6/11/12/21/33 as well as from related and unrelated healthy donors (n=7) were incubated overnight with 100 nM of the Mtphagy dye prior to flow cytometry analysis using the B4 (PE-Vio 770) channel. Shown are the percentage values of the patient (PSMC5 lesions) and control groups (\*\**p*<0.01, Mann-Whitney U test).

To delve deeper into the cellular repercussions of proteasome loss of function, we conducted proteomics investigations on subject-derived T cells. Our findings reveal a strong correlation between proteasome dysfunction and increased expression levels of HLA class I molecules and immune regulatiors, (**Fig. S6, Table S3d**). Furthermore, we observed many enriched proteins involved in amino acid or protein metabolism (FKBP1A, NAPG, LXN, DDAH2, FAH, DPP4, CSTB, GGT3P, AASDHPPT), oxidative stress (GPX4, GSTM1), intracellular transport and signaling (RRAGC, AGA, VPS35L, ARL8A, TBC1D10A, NCALD), regulation of proliferation and cell death (TRIAP1, BRD8, DFFA, CASP6, TRADD), as well as apoliproteins (**Table S3d**).

The massive enrichment of apolipoproteins prompted us to investigate lipid compositions of T cell samples of patients versus healthy controls. Surprisingly, we observed a massively altered distribution of lipids in samples of patients with *PSMC5* variants (**Fig. 6C,D**). Thus, phospholipids of membranes such as phosphatidylserine (PS), phosphatidylethanolamine (PE), phosphatidylcholine (PC), as well as di-(DG) or tri-acylglycerols (TG), were significantly decreased in patients, whereas cholesterolesters were increased by 50%.

The enrichment of mitochondrial proteins (**Fig. S6B**) may point to impaired mitochondrial import or protein quality control. To investigate whether defective proteasomal degradation was accompanied by mitochondrial damage, we monitored mitophagy rates by flow cytometry in S1/6/11/12/21/33, using pH-sensitive MtPhagy dye. Indeed, T cells from individuals with *PSMC5* variants exhibited a higher percentage of lysosomal-targeted mitochondria than controls (**Fig. 6E**). This confirms that proteasome dysfunction is associated with increased degradation of mitochondria in these subjects, indicating mitochondrial damage.

### NDD individuals harboring *PSMC5* variants display sterile type I IFN responses predominantly triggered by the ISR

So far all proteasompathy syndromes (PRAAS and NDD) were linked to dysregulated type I interferon (IFN) signaling irrespective of the gene variants. Indeed, our focused transcriptomic analysis using a NanoString encounter autoimmune gene panel revealed a strongly elevated type I IFN gene signature of five affected individuals compared to healthy controls or relatives (**Fig. 7A**).

**Fig. 7:**
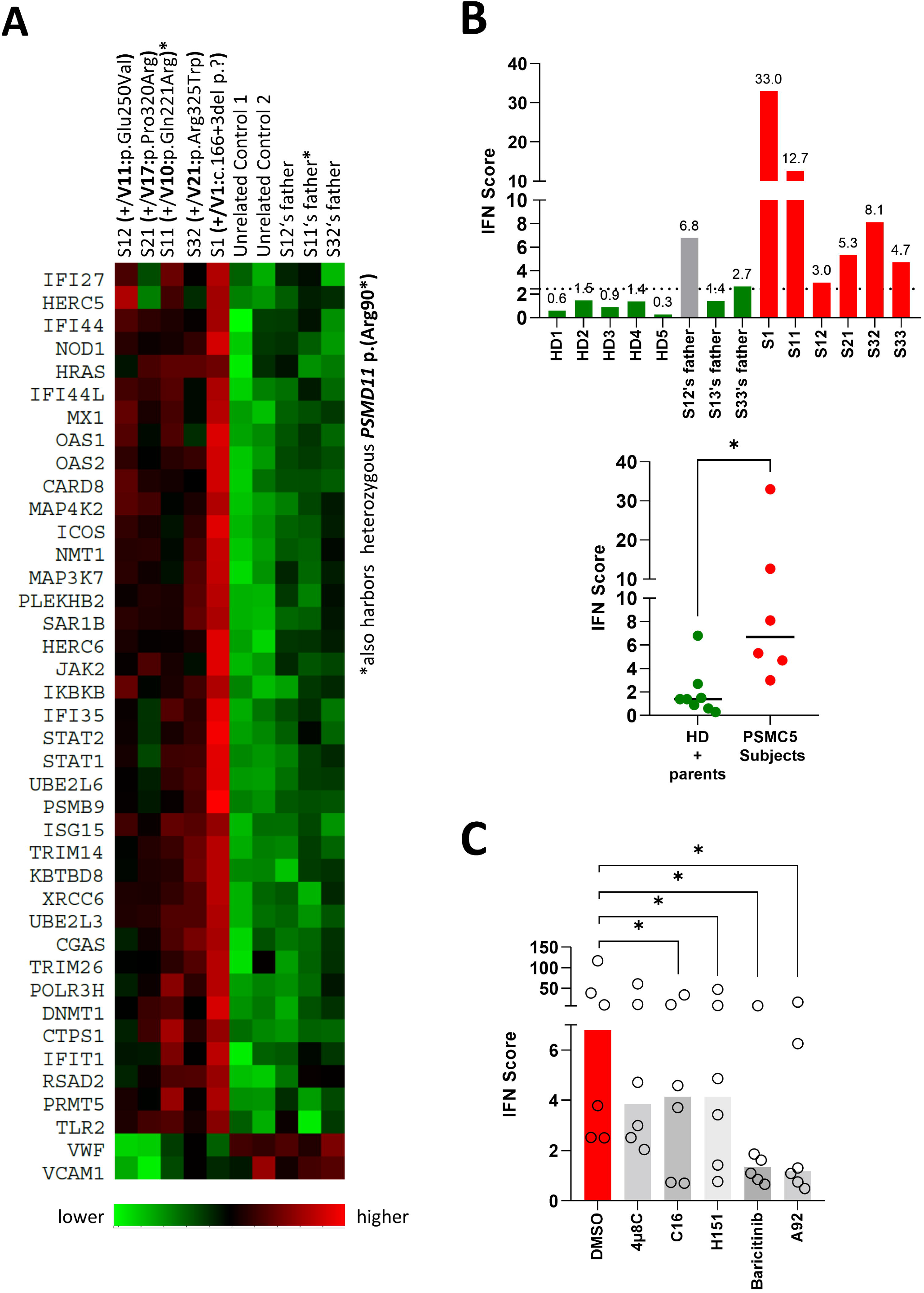
Gene expression analysis reveals a specific gene signature and a spontaneous type I IFN response in T cells from NDD subjects with *PSMC5* heterozygous variants. ***A.*** The heat map indicates the fold-change of expression of immune-related genes between T cell samples from five subjects with NDD (S1/11/12/21/32) and five heathy individuals (probands’ fathers and unrelated donors). Our profiling was based on a panel of 700 immune system genes, using the previously described NanoString nCounter® technology^11^. Differentially expressed genes are hierarchically clustered. Upregulated and downregulated genes are represented in red and green, respectively. Forty genes were differentially expressed, almost all upregulated. The vast majority of them belonged to the large family of type I interferon (IFN)-stimulated genes (ISGs), including in particular the signal transducers STAT1 and STAT2 and members of the ISG15 conjugation machinery (e.g. UBE2L6 and HERC5). ***D*.** A comparative expression analysis of seven ISG (*IFIT1*, *IFI27*, *IFI44*, *IFI44L*, *ISG15*, *MX1* and *RSAD2*) was done by quantitative real-time PCR (see Materials & Methods), as previously described ^11^ between T cells from *PSMC5* subjects S1/11/12/21/32/33, unrelated healthy donors (HD) and subjects’ parents. Type I IFN score was determined for each sample by calculating the median of the normalized fold-change values of the seven ISG relative to one control calibrator. Shown are the IFN scores of each group of donor and parental controls (n=8; left panel) and affected individuals (n=6; right panel). (*p<0.05, Mann-Whitney U test). ***E*.** We evaluated the potential contribution of PKR to the initiation of type I IFN responses in NDD patients with *PSMC5* variants. To this end, T cell from subjects S1/11/12/21/32/33 were exposed to PKR specific inhibitor C16 for 12 h prior to quantification of ISG transcripts by qPCR. T cells expanded from PBMC isolated from NDD subjects were subjected to an 8-hr treatment with a mix including the PKR specific inhibitor C16 (3 µM), as well as DMSO (vehicle), 4µ8C (100 µM), H-151 (2 µM), baricitinib (1 µM) or A92 (10 µM). Then RNAs were extracted and subjected to RT-qPCR for ISG gene expression analysis. Shown are the median values of the IFN scores calculated for each T cell patient sample (n=6) under different treatment conditions (\**p*<0.05, Mann-Whitney U test).

To further confirm the association between *PSMC5* variants and a type I IFN gene signature, we calculated the IFN scores of T cells from S1/11/12/21/32/33 and unaffected controls. These scores wer based on the transcript expression of seven IFN-stimulated genes (ISGs: *IFIT1*, *IFI27*, *IFI44*, *IFI44L*, *ISG15*, *MX1* and *RSAD2*). All affected individuals showed elevated expression of the ISGs compared to controls (**Fig. S7**), resulting in high IFN scores (**Fig. 7B**) and the activation of type I IFN signaling. Intriguingly, the father of S11, carrying the heterozygous *PSMD11* variant p.(Arg90*) (**Table S2**), exhibited a positive type I IFN score (**Fig. 7B**), suggesting that proteasome loss-of-function variants consistently lead to type I IFN gene signatures, even in mildly affected subjects.

Taking into account the established role of protein kinase R (PKR) in initiating sterile autoinflammatory response to proteasome dysfunction^18^, we assessed its contribution to the initiation of type I IFN responses in T cells from subjects with *PSMC5* variant by using C16, a specific PKR inhibitor. The IFN scores from T cells treated with C16 were significantly lower in T cells treated with C16 than in the same T cells exposed to DMSO (controls), confirming PKR’s role in spontaneous ISG induction in this disorder (**Figs. 7C,S8**). A comparable reduction was observed upon H-151 treatment (**Figs. 7C,S8**), underscoring an additional contribution from the cGAS-STING pathway, which senses host-derived DNA in this process. Of note, the IFN scores exhibited a dramatica decrease upon treatment with JAK inhibitor baricitinib (**Figs. 7C, S8**), confirming the autocrine/paracrine nature of this phenomenon. Conversely, blocking the UPR with the IRE1 inhibitor 4µ8C did not yield any significant effects (**Figs. S8**). However, GCN2 inhibition with A92 led to a significant reduction in IFN scores for all subjects, emphasizing the key role of ISR in the generation of type I IFN.

## Discussion

Rare syndromic NDDs caused by *PSMC5* loss-of-function highlight the vulnerability of genes encoding proteasome subunits to genomic lesions, leading to a diverse range of severe neurological phenotypes. *PSMC5* now joins a growing list of proteasome subunit genes linked to various central nervous system (CNS) disorders^9-12^, underscoring the crucial role of the proteasome in development. This has been demonstrated in studies involving plants and mice, where proteasome disruption led to severe organ defects or non-viability^19,20^.

While *PSMC5* is expressed ubiquitously, phenotyping points to a predominant neurodevelopmental phenotype in all affected individuals. The connection between *PSMC5* and NDDs had already been strongly suggested by a large international cohort study, which stressed a significant enrichment of *de novo* variants in this gene^21^. This aligns with the described critical role of PSMC5/Rpt6 in neuronal function, particularly in regulating synaptic function in hippocampal neurons through CAMKIIα-mediated phosphorylation^22,23^. However, the impact of *PSMC5* loss of function extends beyond the CNS, affecting various physiological systems to varying degrees among affected individuals (**Fig. 2A, Tables 2,S2**).

*In vitro* functional investigations on T cells with *PSMC5* variants highlighted a characteristic cellular phenotype consisting of proteasome loss-of-function (**Figs. 5,6**), increased mitophagy (**Figs. 6C,S6B**), and a type I IFN gene signature (**Figs. 7B,S7**). The latter feature bears resemblance to what is observed in individuals with chronic atypical neutrophilic dermatosis with lipodystrophy and elevated temperature (CANDLE, also known as proteasome-associated autoinflammatory syndromes, PRAAS), which are the first described monogenic disorders associated with proteasome gene variants^24^. Nevertheless, unlike individuals with CANDLE/PRAAS, who have severe and typical cutaneous and systemic inflammatory symptoms^24^, the individuals with *PSMC5* variants in this series do not display striking signs of autoinflammation, although pathogenesis mechanisms partially overlap in proteasome impairment, activation of proteotoxic stress and dysregulation of type I IFN signaling. Similar to the Aicardi-Goutières-Syndrome, a prototypic type I interferonopathy with CNS manifestation, developmental defects such as growth retardation also have been observed in patients with CANDLE/PRAAS^24^.

Although it remains unclear whether dysregulated type I IFN production actively contributes to the development of NDD phenotypes, it is noteworthy that two potential markers of the disorders described here, PKR and GCN2—both sensors of the ISR—belong to the eIF2α kinase (EIF2AK) family. This kinase family has already been associated with NDD through its members *EIF2AK1* and *EIF2AK2*^25^. Interestingly, in mammals, GCN2, a regulator of neurogenesis known for its inhibitory effect on spontaneous neuritogenesis, also plays a significant role in behavioral control and memory consolidation^26^. In turn, PKR is involved in the pathogenesis of neurodegenerative disorders, notably Alzheimer’s disease^27^. In this context, our results hold great promise, as type I IFN responses in T cells harboring *PSMC5* variants could be reduced using inhibitors of PKR, GCN2, and JAK (**Figs. 7C,S8**). These findings merit further investigation in clarifying the role of inflammation on neurogenesis in the *PSMC5*-associated disorder.

*PSMC5* variants notably disrupt lipid homeostasis: (i) in S6 and S11, we noted an enrichment of apolipoproteins APOA1, APOA2, APOE, and APOC3 **(Fig. S6)**, whose degradation partially rely on proteasomes^28,29^; and (ii) in S1/6/11/12/19/21/33’s cells, we measured reduced amounts of triglycerides, diglycerides and glycerophospholipids, while their pools of cholesterol ester were increased **(Fig. S6D)**. Perturbations of lipid homeostatis were noted in CANDLE/PRAAS subjects, typically suffering from lipodystrophy^24^. Furthermore, lipid remodeling has been associated with other neurodevelopmental pathologies, including ASD^30^ and ADHD^31^.

To emphasize the neuronal implications of *PSMC5* variants, we employed both hippocampal cell and *Drosophila melanogaster* models. Assays in primary hippocampal neurons suggested that LoF variants impair the neuritogenic role of *Psmc5* (**Fig. 4**), Additionally, the *Drosophila Psmc5*-knockdown (KD) model demonstrated the influence of *Psmc5* loss on cognitive performance. Our assays showed that *Psmc5* KD impaired reversal learning without compromising memory (**Fig. 3A,B**). These findings echo observations from the paralogous gene *PSMC3*, whose KD produces similar effects in flies^11^. Additionally, parallels emerge from studies showcasing behavioral deficits in male rats with reduced proteasomal activity during memory recovery^32^. Interestingly, mouse investigations suggested the involvement of *Psmc5* in molecular and behavioral plasticity, particularly in response to stimuli like cocaine^33^. These studies demonstrated that Psmc5/Rpt6, when forming a complex with ΔFosB and other proteins in the nucleus accumbens, regulates chromatin remodeling and gene expression. This observation gains significance when considering the enriched presence of dopaminergic neurons in the midbrain region, which is pivotal in behavioral flexibility^34^. In addition to its connection with ΔFosB, PSMC5/Rpt6 was found to physically interact with type B GABA (GABAB) receptors, modulating their presence on cell surfaces and subsequently affecting neuronal activity^35^. This suggests a potential link between *PSMC5* variants and GABAB signaling, which is a key element in behavioral flexibility^36^ and impaired in ASD and ADHD^37^.

In the last experimental model involving rat hippocampal cells, *Psmc5* KD induced a lowered E/I ratio (**Fig. 3G-J**). This phenomenon has been associated with modified cognitive function in humans, affecting decision-making negatively^38^. The imbalance in E/I ratio is implicated in conditions such as ASD and schizophrenia, where it disrupts brain neural circuits and leadds to a decline in cognitive abilities^39^.

Collectively, our findings obtained from the various experimental models of the PSMC5-associated NDD highlight parallels with aging and neurodegeneration. T cells derived from individuals with *PSMC5* variants exhibit characteristics reminiscent of senescent cells, including decreased proteasomal activity in T cells indicated by the accumulation of polyubiquitinated proteins (**Fig. 6A,B**). Proteasomal activity is known to decline with age in humans^40^, and its beneficial impact on lifespan has been extensively documented in human bone marrow multipotent stromal cells^41^ or *Drosophila*^42^ models. Also consistent with observations in T cells (**Figs. 6C,7,S6,S7**), age-related cellular traits comprise impaired mitophagy^43^, type I IFN response^44^, and elevated levels of APOA2^45^. The elevated prelamin A (LMNA) levels observed in affected individuals (**Fig. S6C**) mirror the excess LMNA resulting from mTOR activation reported in a rat progeria model, where it correlated with premature aging and compromised autophagy^46^. These observations could potentially clarify the usage of phosphorylated Psmc5 (Rpt6) and the accumulation of K48-linked polyubiquitinated proteins as markers to monitor proteasomal activity in the aging brain of another rat model^32^. These findings raise questions about whether *PSMC5* variants induce premature aging in the affected individuals and whether they play a role in neurodegenerative diseases. Indeed, reduced proteasome activity significantly contributes to the onset and progression of age-related neurodegenerative diseases, such as Alzheimer, Parkinson or Huntington disease^40^. Interestingly, the conditional inactivation of *Psmc1*, a paralog of *Psmc5*, successfully recapitulates Parkinson disease phenotype in both mice^47^ and *Drosophila*^48^. Despite the average pediatric age of the *PSMC5* individuals (9.1 years) and the limited follow-up on disease progression, it is noteworthy that three cases of developmental regression have been obserrved in the series. Additionally, our findings inform the neuronal or animal models also concur with the hypothesis linking *PSMC5* variants to senescence and age-related dementias. Impaired neuritogenesis, for instance, was reported for instance in an iPSC-derived neuronal model of Parkinson’s disease^49^, cognitive inflexibility was observed in individuals with Parkinson’s and Alzheimer’s diseases^37^, and imbalance in E/I ratio was similarly noticed in individuals with Alzheimer’s disease^50^.

To conclude, the combined inputs of T cells, neuronal models, animal studies, and cognitive paradigms contribute to a multi-dimensional understanding of the pivotal role played by PSMC5/Rpt6 in neurodevelopment and neuronal function. Our data reveal major overlaps in the molecular pathogenesis of NDD stemming from *PSMC5*, *PSMC3* and *PSMD12* variants which are collectively characterized by protein aggregation (**Fig. 6A, B**), increased mitophagy (**Fig. 6E**) and activation of the ISR leading to sustained type I IFN production (**Figs. S7,8**)^10,11^, which, very engagingly, can be mitigated by the use of inhibitors targeting PKR, GCN2, or JAK.

These findings not only enhance our understaning of the underlying mechanisms of neurodevelopmental proteasomopathies, but also lay the foundation for novel diagnostic procedures and potential therapeutic strategies. While the primary focus will be on individuals with these specific proteasome-related disorders, the remarkable similarities observed between the cellular characteristics of individuals with *PSMC5* variants and those with neurodegenerative diseases suggest that these therapeutic strategies could eventually benefit a broader population affected by neurodegenerative conditions.

## Supporting information

Supplemental notes and figures - updated

Supplemental Table 1 - updated

## Web resources

Combined Annotation Dependent Depletion (CADD), https://cadd.gs.washington.edu/

dbSNP, http://www.ncbi.nlm.nih.gov/projects/SNP/

DIOPT, https://www.flyrnai.org/cgi-bin/DRSC_orthologs.pl

GeneMatcher, https://genematcher.org/

gnomAD, http://gnomad.broadinstitute.org/

InterVar, https://wintervar.wglab.org/

Metadome, https://stuart.radboudumc.nl/metadome/

Missense Tolerance Ratio Gene Viewer, http://mtr-viewer.mdhs.unimelb.edu.au/

MobiDetails, https://mobidetails.iurc.montp.inserm.fr/MD/

OMIM, http://www.omim.org/

PRIDE, http://www.ebi.ac.uk/pride/

RCSB Protein Data Bank, https://www.rcsb.org/

UCSF ChimeraX, https://www.rbvi.ucsf.edu/chimerax/

## Data availability

Mass spectrometry proteomics data are available via ProteomeXchange with identifier PXD048558. Lipidomics data have deposited in a repository through the Metabolomics Workbench; the identifier will be communicated once attributed.

## Acknowledgments

This study is part of the GEM-EXCELL, a network of excellence in genetics and genomics embedded in the French network of University Hospitals HUGO (‘Hôpitaux Universitaires du Grand Ouest’). This work has been carried out within the framework of the FHU GenOMedS thanks to the support of the Health cooperation group of University Hospitals of the Great West (GCS HUGO) and the National Alliance for Life Sciences and Health (Aviesan). The Deciphering Developmental Disorders (DDD) study presents independent research commissioned by the Health Innovation Challenge Fund (grant number HICF-1009-003). IDB, MDD’A and MKL are contributors to the Care for Rare Solve study led by the Care for Rare Canada Consortium, Children’s Hospital of Eastern Ontario Research Institute, Ottawa, ON, Canada. The authors would like to express their sincere gratitude to Tayyaba Khan and Cherith Somerville for their efforts in clinical data collection and management.

