## Supplemental notes and figures - updated for "Unveiling the crucial neuronal role of the proteasomal ATPase subunit gene *PSMC5* in neurodevelopmental proteasomopathies"

**Supplementary information**

**Supplementary material and methods**

**Plasmid construction and transfection**

The cDNA for *PSMC5* coding wild-type PSMC5 (i.e. Rpt6) full-length protein was amplified by RT-PCR from total RNA isolated from HeLa cells and cloned into the pcDNA3.1/Zeo(+) (Invitrogen) using the *Kpn* I and *Xho* I restriction sites. For N-terminal tagging, oligonucleotides encoding a tandem HA epitope (GYPYDVPDYAMGGYPYDVPYAGT) were constructed, annealed and inserted in frame into the pcDNA3.1/Zeo(+) expression vector containing *PSMC5* to generate a HA-PSMC5 (i.e. HA-Rpt6) fusion protein. Thirteen cDNA variant constructs were then prepared according to mRNA sequence NM_002805.6, and contained respectively the variants c.414G>C p.(Pro183Leu); c.601C>T p.(Arg201Trp); c.605C>T p.(Ala202Val); c.620C>T p.(Thr207Met); c.647G>A p.(Gly216Asp); c.662A>G p.(Gln221Arg); c.749A>T p.(Glu250Val); c.773G>A p.(Arg258Gln); c.959C>G p.(Pro320Arg); c.959C>A p.(Pro320His); c.973C>T p.(Arg325Trp); c.1103T>C p.(Met368Thr); c.1182_1183del p.(Asp394Glufs*2). These *PSMC5* variants were introduced into the pcDNA3.1/HA-*PSMC5* plasmid by site-directed mutagenesis, using appropriate primer pairs designed with the Quick-Change Primer Design software from Agilent. The JetPRIME reagent (Polyplus-transfection® SA) was employed to transfect SH-SY5Y cells according to the manufacturer’s instructions.

**Proteomic analyses of human T cells**

*Preparation of protein samples.* Protein was extracted from primary T cells by five cycles of freezing (liquid nitrogen) and thawing (30^o^C, 1,400 rpm) in 8 M urea/ 2 M thiourea. Cell debris and insoluble material was separated by centrifugation (16,000 × g, 1 h at 20 C). Protein content was determined with a Bradford assay (Biorad, Munich, Germany).

*Sample preparation* *for mass spectrometry*. Four µg of total protein from each sample were reduced (2.5 mM DTT ultrapure, Invitrogen, for 15 min at 37 °C) and alkylated (10 mM iodacetamide, Sigma Aldrich, for 30 min at 37°C). Benzonase (Novagen, Merck Millipore, Darmstadt Germany) 0.6 U/µg protein) was applied for nucleic acid degradation before digestion with trypsin (Promega, Madison, WI, USA) at an enzyme to protein ratio 1:25 over night at 37 °C. The tryptic digestion was stopped by adding acetic acid (final concentration 1%) followed by desalting using ZipTip-µC18 tips (Merck Millipore, Darmstadt, Germany). Eluted peptides were concentrated by evaporation under vacuum and subsequently resolved in 0.1% acetic acid, 2% acetonitrile (ACN) containing HRM/iRT peptides (Biognosys, Zurich, Switzerland) according to manufacturer’s recommendation. For the generation of a spectral library a protein pool of equal protein amounts (10 µg) of each sample was prepared as described above. Resulting peptides were desalted (SepPak, Waters, Eschborn, Germany) and fractionated by strong cation exchange chromatography (PolySulfoethyl A™ column, 150 x 1mm, 5µm, 200A, Poly LC Inc., Columbia, ML, USA). Peptide fractions (n=17) were purified for mass spectrometric analyses as described above.

*Mass Spectrometry Measurements*. Mass spectrometric (MS) data was recorded on a QExactive HF mass spectrometer (Thermo Electron, Bremen, Germany). Before MS data acquisition tryptic peptides were separated by reverse phase chromatography (Accucore 150-C18, 25 cm x 75 μm, 2,6 μm C18, 150 Å) using an Ultimate 3000 nano-LC system (both Thermo Scientific, Waltham, MA, USA) at a constant temperature of 40°C and a flow rate of 300 nL/min. To design a spectral library, MS/MS peptides were separated by 120 min linear gradients with increasing acetonitrile concentration from 5 to 25 % in 0.1 % acetic acid. Data were recorded in data dependent mode (DDA). The acquisition of MS data for relative quantitation was performed in data independent mode (DIA) after peptide pre-fractionation at chromatographic conditions described above. For further details to the instrumental setup and the parameters for LC-MS/MS analysis in DDA and DIA mode see supplemental **Tables S3a** and **S3b**.

*Data analysis.* Proteins were identified using Spectronaut^TM^ Pulsar 13.9 software (Biognosys AG) against a spectral library generated from data-dependent acquisition measurements of a SCX-fractionated peptide pool. The spectral library construction by Spectronaut was based on a database search using a human protein database (Uniprot vs 03_2019, 20404 entries) (**Table S3c**). The generation of the ion library in Spectronaut^TM^ v13.9.191106.43655 resulted in a constructed library consisting of 920,617 fragments, 67,323 peptides and 6717 protein groups. The Spectronaut DIA-MS analysis was carried out as described previously^1^ with project specific modifications (**Table S3c**). Peptides were assigned to protein groups and protein inference was resolved by the automatic workflow implemented in Spectronaut. Only proteins with at least two identified peptides were considered for further analyses. Data analysis was performed with an in-house R-pipeline. Data was median normalized on ion level. Statistical analysis was carried out on peptide level using the algorithm ROPECA^2^. Peptides with oxidized methionine were not included in the quantitative analysis. Binary differences have been identified by application of a moderate paired t-test^3^. Family was used as pairing variable. Multiple test correction was performed according to Benjamini-Hochberg. Variance within the data set was visualized by principal component analyses and differences in the protein pattern by Volcano plots (data not shown); for representation of protein intensities Hi3 Peptides were used. Data obtained for the quantification of Hi3 peptides are reported in **Tables S3d** and **S3e**. **Table S3d** reports the separate comparisons between the individual intensities of Hi3 peptides obtained for Subject S6 and her mother, on the one hand, and Subject S11 and her father, on the other hand. **Tables S3e** reports the comparisons between combined intensities of Hi3 peptides from the affected individuals (Mut= Subjects S6 and S11) and combined intensities from their parental controls (CO= Subject S6’s mother and Subject S11’s father).

The mass spectrometry proteomics data have been deposited to the ProteomeXchange Consortium via the PRIDE^4^ partner repository with the dataset identifier PXD048558.

**Functional enrichment analysis from proteomics data**

The statistical analysis was performed on proteins with more than 2 peptides, using intensity data obtained by Hi3 peptides quantification reported in **Table S3e**. All proteins with p-value<0.05 were included in the functional enrichment (**Fig S6A**). The functional analysis was performed using the R package gprofiler2 was used to perform pathway enrichment analyzes (DB: GO:BP,KEGG, REAC and WP) with gene symbols as input. The following options were selected: correction_method=fdr, user_threshold=0.05.

A Fisher's exact test was performed to determine if the significant proteins were enriched for specific biological terms of interest: The input data consisted of a list of proteins that were differentially expressed between patients and controls (p-value < 0.05). (i) The reference database containing information about mitochondrial genes was the MitoCarta3.0 human inventory implemented by the Broad Institute (<https://www.broadinstitute.org/files/shared/metabolism/mitocarta/human.mitocarta3.0.html>); it consists in ‘a collection of 1136 nuclear and mtDNA genes encoding proteins with strong support of mitochondrial localization’^5^. (ii) The aging gene list was obtained by searching the query ('longevity' OR 'lifespan' OR 'CLS' OR 'chronological life span' OR 'ageing') AND 'Homo sapiens'[porgn:__txid9606] on https://www.ncbi.nlm.nih.gov/gene. The enrichment test assessed whether certain biological terms were overrepresented in the input list compared to what would be expected by chance, using a Fisher's exact test (**Table S3f**).

**Supplementary results**

**Three-dimensional (3D) structural analysis predicts that missense *PSMC5* variants affect proteasome function in various ways**

Most *PSMC5* variants affect residues located near neighboring ATPases from the base of the 19S regulatory particle. For instance, Met368 appears to interact with PSMC4/Rpt3 by forming interactions with the hydrophobic moieties of Tyr191, Pro199 and Arg 329 (**Fig. S2D**). Its substitution by a polar threonine, thereby creating a change in the electrostatic environment on the subunit’s interface, might compromise complex formation and the integrity of this part of the 26S proteasome. Furthermore, 6/13 residues play a role in the inter- and intramolecular rearrangements that occur during the transition from substrate-free to substrate-engaged states of the 26S proteasome (**Fig. S2B**), suggesting that alteration of these amino acids could disrupt this phase. For example, Arg201 in PSMC5/Rpt6 forms a polar interaction network with PSMC4/Rpt3 near the ATP-binding pocket (**Fig. S2B**). Glu250 is close to the ATP binding site though not close enough to form a contact with the charged phosphate (**Fig. S2B**). Since ATP binding appears to be essential for proteasome assembly^6^, modifications of amino acids at the ATP binding pocket may affect ATP binding and hydrolyzation and thereby the integrity of the tertiary structure. This would explain the drastic effect of these two variants on *PSMC5* expression. Arg258 forms a contact network with PSMC1/Rpt2 near the substrate binding channel (**Fig. 1B,S2C**). As this variant is well expressed and incorporated in *in vitro* experiments, it stands to reason that it mainly plays a role in substrate processing, although we did not further confirm its effect on proteasome activity in our analysis. In presence of this variant, the chemical identity of the side chain is altered, which could influence the dynamics of the subunit and thus substrate processing. In addition to a change in the physicochemical properties of the side chains, there are some variants that are more likely to affect the flexibility or rigidity of the protein backbone. We identified four variants in which either a proline or a glycine is altered (Pro183Leu, Gly216Asp, Pro320Arg, Pro320His). Glycine typically allows for backbone flexibility, while proline enforces conformational rigidity and often induces turns. Predicting the exact effects of these four variants on protein folding and activity remains challenging. However, since both Pro183 and Pro320 are situated within turns between two secondary structure elements (**Fig. 1B,S3**), it is highly likely that these changes will impact protein folding and incorporation of subunits. Furthermore, Pro320 and Arg325 (Arg325Trp) are part of the P-loop between the core and the AAA+-lid domain. Arg325 forms multiple hydrogen bonds connecting these two domains (**Fig. S3**). Altogether, these findings strongly suggest that substitutions in PSMC5/Rpt6 are likely to have multifaceted effects on proteasome.

***PSMC5* loss-of function is associated with proteomic changes affecting immunity, metabolism and cell proliferation**

Pairwise comparison and hierarchical clustering analysis of proteomics data identified approximately 20 proteins significantly enriched in subjects S6 and S11 compared to their parents **(Fig. S6A)**: (i) players of the innate and adaptive immune responses, such as APOBEC3G or HLA class I (HLA-A) and II (HLA-DRB1) molecules, which are typically induced by type I and II IFN, respectively^7^; (ii) a set of apolipoproteins (APOA1, APOE, APOA2, APOC1, APOC3, APOA4, APOBR and APOL3) (iii) proteasome subunit PSMB7 (i.e. β2), suggesting that T cells with *PSMC5* variants attempt to restore protein homeostasis by inducing the synthesis of new proteasome complexes, as previously described^8^; (iv) prelamin A (LMNA); (v) proteins related to innate immunity (PPP1R11), (vi) amino acids (FAH), or (vii) glutathione (GSTM1). Note that some of these proteins were also relatively abundant in S10’s father –exhibiting moderate NDD- who transmitted alteration *PSMD11* NM_002815.3:c.268C>T p.(Arg90*) to his daughter, confirming the likely pathogenicity of this second proteasome variant. Finally, a group of proteins were consistently downregulated across NDD patients, most of them regulators of cell proliferation, such as CDC123, REPS1 and AAGAB **(Fig. S6A)**. Altogether, these investigations uncovered a series of candidate biomarkers for NDD caused by proteasome defects which themselves reflect profound remodeling of immune responses and lipid metabolism.

A Fisher's exact test was performed to determine if the significant proteins were enriched for specific biological terms of related either to mitochondria or aging. According to the lists provided for both biological terms, amongst all the proteins that could be tested by proteomic analysis, 4 of the 55 proteins found significantly deregulated (p-value<0.05) are linked to mitochondria (NIT1, MTHFD2, GPX4, TRIAP1 on volcano plot in **Fig. S6B**). However, the p-value of hypergeometric test related to the whole list of mitochondrial genes is not significant (p-value = 0.9). By comparison, 6 of the 55 significantly deregulated proteins are related (LMNA, APOE, HLA-DRB1, HLA-DQA1, APOC3, GSTM1 on volcano plot in **Fig. S6C**), with a significant p value of the hypergeometric test related to the list of aging genes (p-value = 0.007).

**Supplementary figures and tables**

**Table S1:** Predictions and functional tests for the PSMC5 variants identified in the affected individuals included in the study.

**Table S2**: Clinical features of the subjects with *PSMC5* variants and indels.

**Table S2a**: Detailed clinical features of the subjects with *PSMC5* variants and indels.

**Table S2b**: Clinical features described by Human Phenotype Ontology (HPO) terms across the patient cohort.

**Table S2c**: Individual lists of symptoms and signs observed in each affected subjects of the study.

**Table S3.** Proteomics analyses.

**Table S3a.** LC-MS/MS parameter (data dependent mode, spectral library).

**Table S3b.** LC-MS/MS parameter (data independent mode; quantitative data).

**Table S3c.** Peptide and protein identification parameters in Spectronaut.

**Table S3d.** Results of proteome-wide analysis indicated by Hi3 peptide intensity data: comparison at the family level between individual data from affected individuals S6 and S11 and individual data from their parental controls.

**Tables S3e.** Results of proteome-wide analysis indicated by Hi3 peptide intensity data: comparison between combined data from affected individuals S6 and S11 and combined data from their parental controls.

**Tables S3f.** List of genes related to aging used for functional enrichment analysis from proteomics data.

**Fig. S1**: Evolutionary conservation of the PSMC5/Rpt6 residues subjected to missense mutations associated with NDD.

**Fig. S2:** Structural analysis of *PSMC5* variants shows that most missense variants are located close to neighboring ATPases PSMC1/Rpt2 and PSMC4/Rpt3.

**Fig. S3:** Structural analysis of PSMC5 variants: detailed view of the Rpt6 region subject to recurrent variants.

**Fig. S4**: Distribution of mean pairwise distance between facial features of subjects with *PSMC5* variants.

**Fig. S5**: *PSMC5* variants exhibit diverse effects on PSMC5/Rpt6 protein turnover and assembly into mature 26S proteasome complexes in SHSY5Y neuroblastoma cells.

**Fig. S6:** Proteome profiling of T cells from NDD subjects identifies candidate biomarkers for proteasome dysfunction.

**Fig. S7**: T cells from NDD subjects with *PSMC5* heterozygous variants exhibit a strong upregulation of ISG.

**Fig. S8**: Sterile type I interferon (IFN) responses triggered in NDD subjects with *PSMC5* heterozygous variants predominantly rely on the ISR.

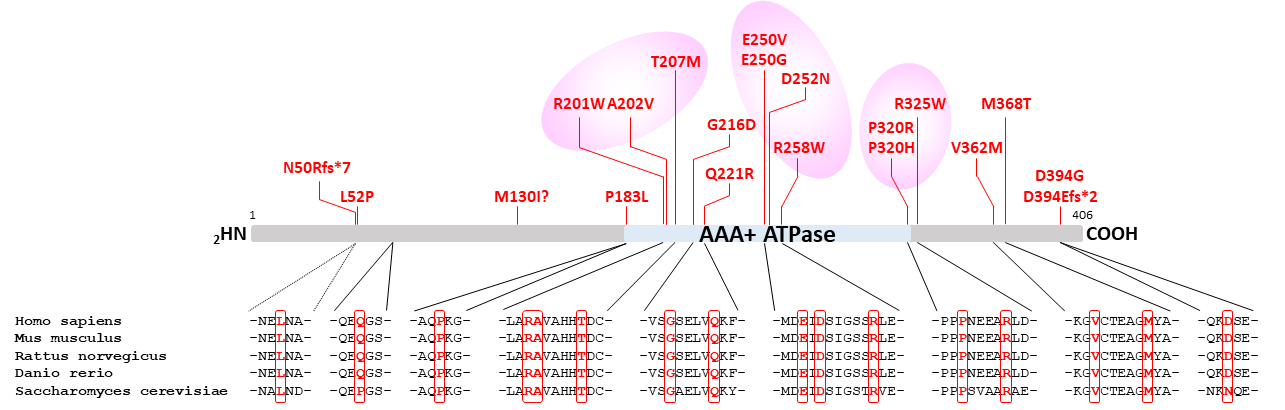

**Fig. S1: Evolutionary conservation of the PSMC5/Rpt6 residues subjected to missense variants associated with NDD**. Alignment of the primary structures of human, mouse, rat, zebrafish and yeast PSMC5/RPT6 showing that all affected residues are evolutionary well conserved across species.

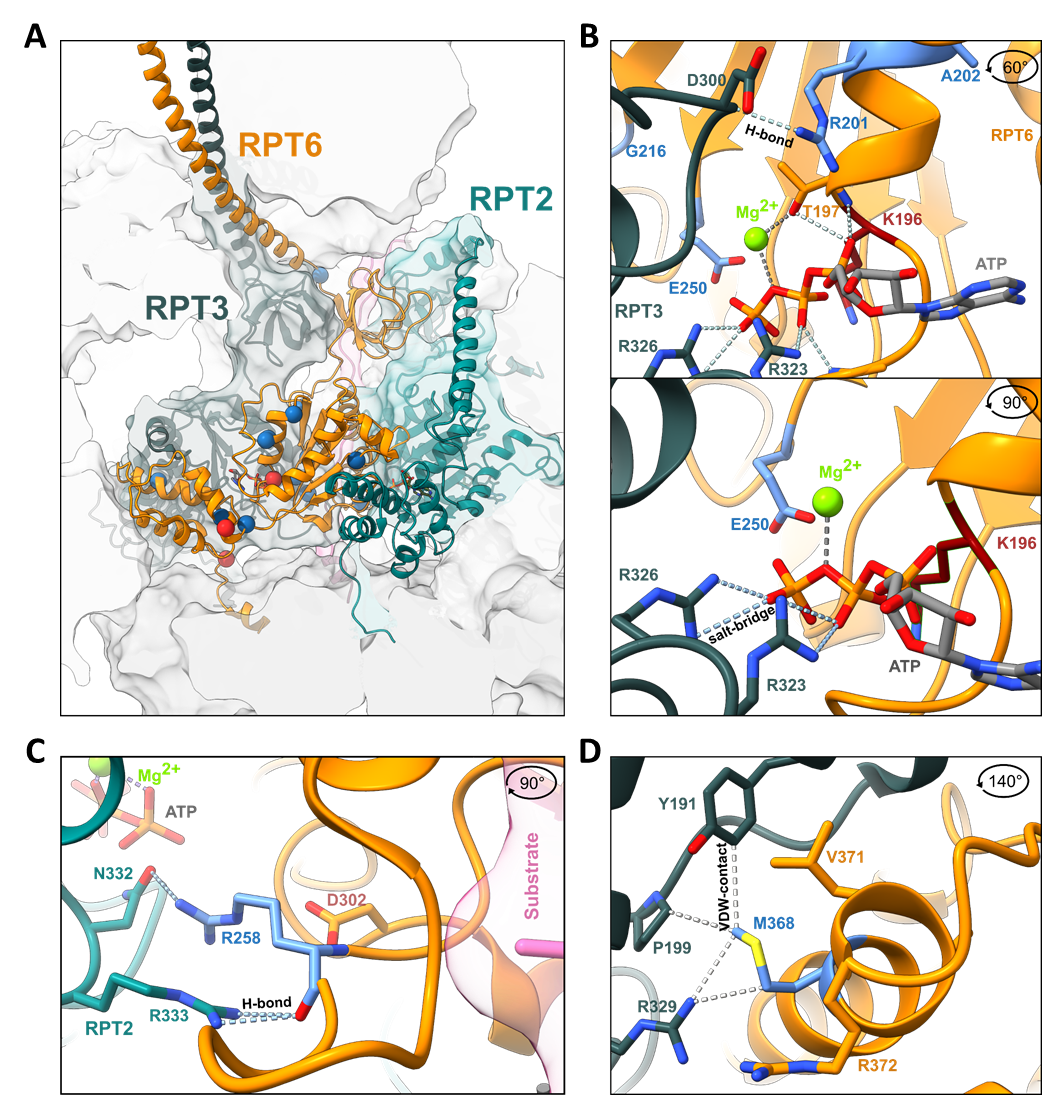

**Fig. S2: Structural analysis of *PSMC5* variants shows that most missense variants are located close to neighboring ATPases PSMC1/Rpt2 and PSMC4/Rpt3. A.** General overview. The variants or interest (blue spheres for missense variants and red spheres for frameshift variants) are visualized within the structure of Rpt6/PSMC5 (orange) in the substrate engaged Cryo-EM structure 6MSK and the substrate free structure 7W37^9,10^. They are primarily located in the lower ATPase domain close to interfaces to the Rpt2/PSMC1 (dark green), Rpt3/PSMC4 (light green), and the substrate (pink). **B-D.** In a significant number of the variants identified in affected individuals, subunit interfaces are modified, as illustrated by the (**B**) detailed view of Glu250 (E250; p.Glu250Gly, p.Glu250Val), Arg201 (R201; p.Arg201TRp) and Lys196 (K196; p.Lys196Argfs*29) which forms bonds with the residues Arg323 (R323) and Arg326 (R326) of Rpt3/PSMC4, (**C**) detailed view of Arg258 (R258, p.Arg258Trp, p.Arg258Gln) which forms hydrogen bonds with Asp300 (D300), Arg233 (R233) and Asn332 (N332) of Rpt2/PSMC1, respectively, and (**D**) and detailed view of Met368 (M368; p.Met368Thr) which forms hydrophobic interactions with residues in the small subdomain of the Rpt3 ATPase domain. Hydrophobic interactions are indicated as grey dashed lines.

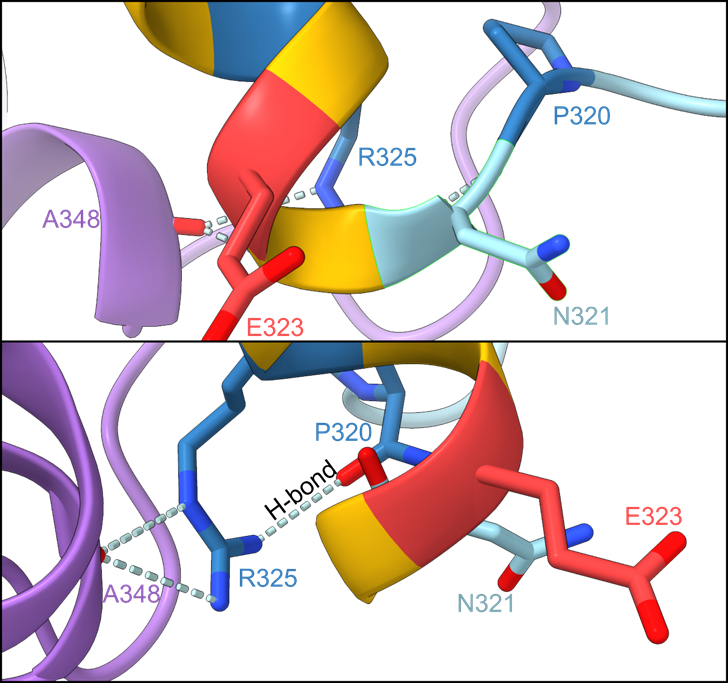

**Fig. S3: Structural analysis of *PSMC5* variants: detailed view of the Rpt6 region subject to recurrent variants.** Detailed view of the residues Pro320 (P320; p.Pro320Arg, p.Pro320His), Arg325 (R325; p.Arg325Trp) and Glu323 (E323; predicted p.Glu323del).

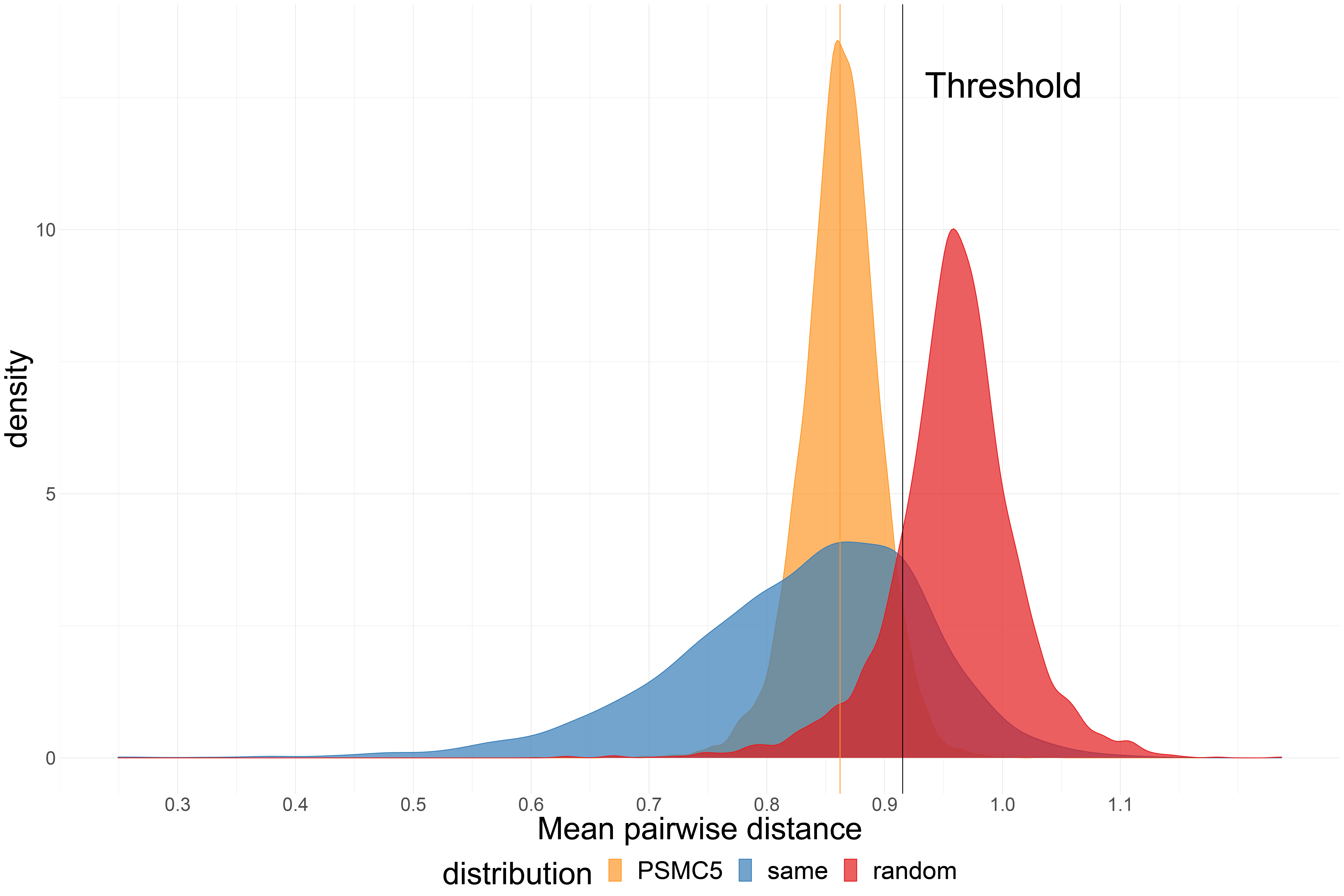

**Fig. S4: Distribution of mean pairwise distance between facial features of subjects with *PSMC5* variants.** It shows three distributions: *PSMC5*, the random selection from the subjects with 328 disorders, and the selection with the same disorder. Black vertical line is the threshold that classifies whether it is the same disorder or random selection. 95% of *PSMC5* distribution are below the threshold.

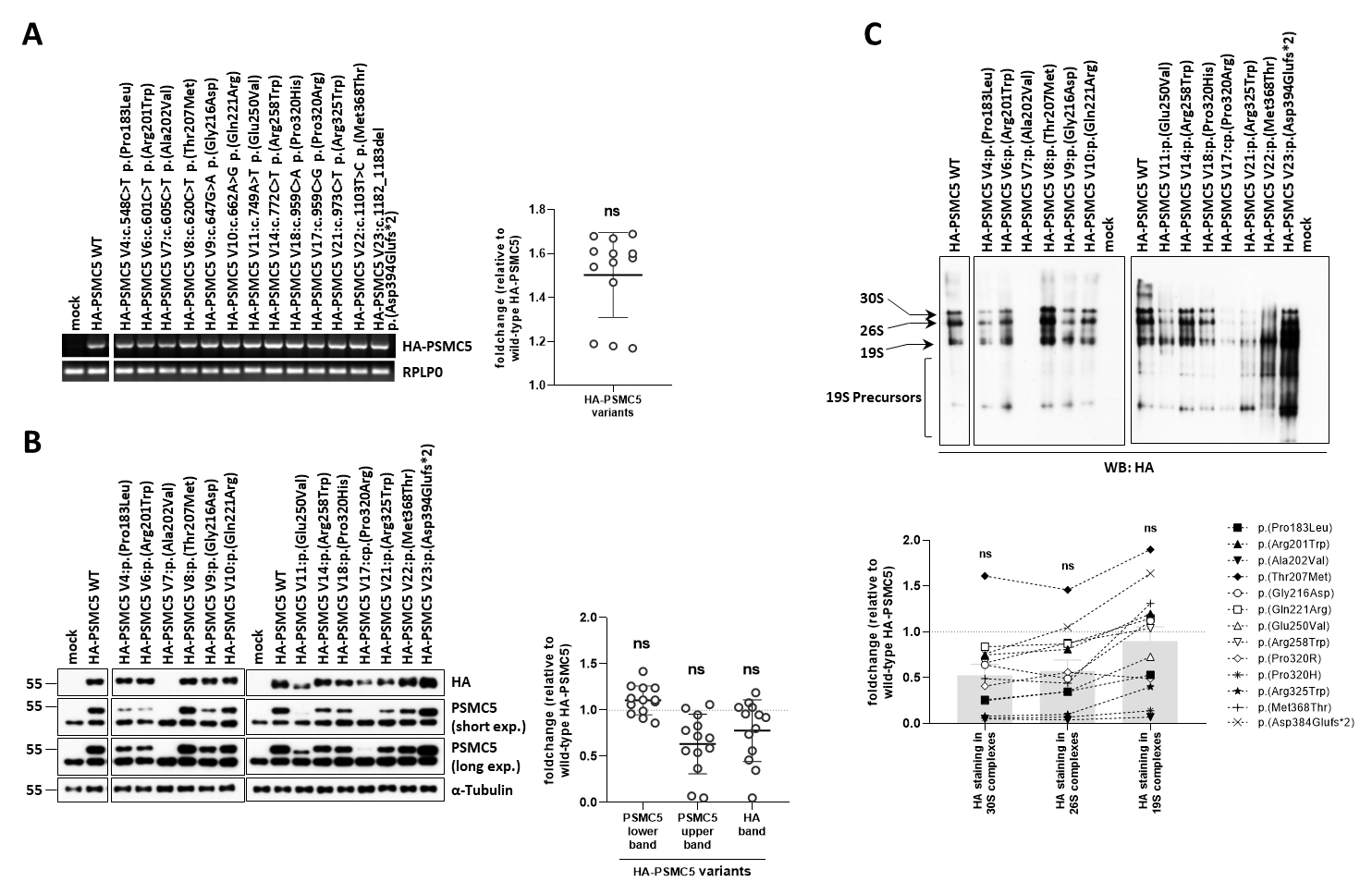

**Fig. S5: *PSMC5* variants exhibit diverse effects on PSMC5/Rpt6 protein turnover and assembly into mature 26S proteasome complexes in SHSY5Y neuroblastoma cells. *A*.** SHSY5Y cells transfected for 24h with wild-type and mutant constructs underwent semi-quantitative RT-PCR analysis to assess *PSMC5* gene expression. Endogenous *PSMC5* amplification was prevented with a BGH reverse primer binding to the polyadenylation signal of the expression vector used for *PSMC5* overexpression. Right panel: HA-PSMC5 PCR bands were quantified by densitometry analysis and normalized to those of RPLP0. Data are presented as normalized foldchange mean values ± SD of the HA-*PSMC5* variants (n=13) over their wild-type counterparts whose densitometric values were set to 1 (grid line). Normalization utilized amplification of the RPLP0 house-keeping gene for equal loading. ***B*.** SDS-PAGE/western blot analysis of 10 µg RIPA-extracted whole-cell lysates expressing N-terminally HA-tagged PSMC5 mutants detected HA and PSMC5/Rpt6 using specific antibodies, and normalizing by GAPDH. Dual time exposures revealed varied effects of *PSMC5* variants on steady-state PSMC5/Rpt6 expression levels evidenced by relative intensities of upper immunoreactive bands, corresponding to HA-tagged PSMC5/Rpt6 full-length protein products of overexpressed *PSMC5* constructs. Right panel: the HA and PSMC5 immunoreactive bands were quantified by densitometry and normalized to those of α-tubulin for each sample. Data are presented as normalized fold change mean values ±SD of PSMC5 (upper and lower bands) and HA obtained with HA-PSMC5 variants (n=13) versus their wild-type counterparts whose densitometric measurements were set to 1 (grid line). ***C.*** Native-PAGE/western blot analysis of 20 µg TSDG-extracted lysates obtained under non-denaturing conditions from the variant and wild-type SHSY5Y cell lines evaluated the incorporation of 13 variant PSMC5/Rpt5 protein products into mature 26S proteasome complexes. Membranes were probed with a monoclonal antibody directed against the HA epitope. Arrows indicated migration of PSMC5-containing 30S, 26S and free 19S complexes, while a bracket marked a set of faster-migrating 19S assembly intermediates. The efficient incorporation of wild-type HA-tagged PSMC5/Rpt5 into unbound 19S regulatory particle positions, single-capped (19S-20S or 26S) and double-capped (19S-20S-19S or 30S) proteasomes, was evidenced by three migrating HA species. The efficient incorporation of wild-type HA-tagged PSMC5/Rpt5 into 19S, 26S and 30S complexes was evidenced by three HA species migrating at the positions of unbound 19S regulatory particle as well as single- (i.e. 19S-20S or 26S) and double-capped (i.e. 19S-20S-19S or 30S) proteasomes. Lower panel: the HA immunoreactive bands in the 30S, 26S and 19S complexes were quantified by densitometry and presented as fold changes for each HA-PSMC5 variant versus wild-type HA-PSMC5 controls whose densitometric values were set to 1 (grid line). Columns represent the foldchange mean values ± SD of the 13 investigated HA-PSMC5 variants, as indicated.

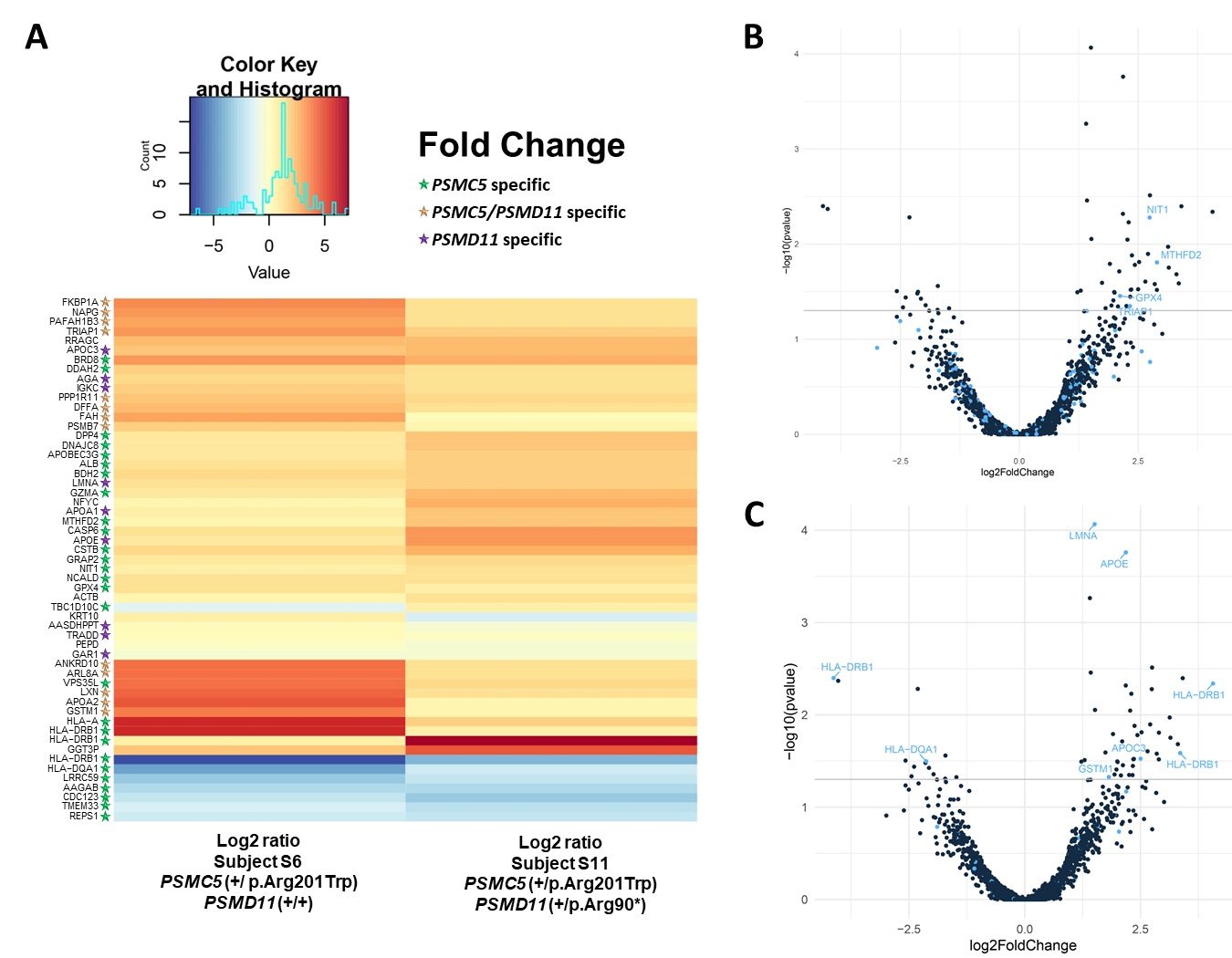

**Fig. S6: Proteome profiling of T cells from NDD subjects identifies candidate biomarkers for proteasome dysfunction.** To decode the cellular effects of *PSMC5* loss-of-function causing NDD, the proteomes of T cells from subjects S6 and S11 were next analyzed using mass spectrometry-based proteomics and compared to those of unaffected parents (mother and father, respectively). **A.** Heatmap of protein abundance pattern showed significantly altered proteins in S6 and S11 compared to parental controls, as indicated (p-value <0.05). Brackets indicate sets of proteins specifically altered in cells carrying *PSMC5*, *PSMD11* or a combination of *PSMC5* and *PSMD11* variants, as indicated. **B, C.** Volcano plot of deregulated expression of genes in enrichment pathways. **(B)** The blue points indicate the genes encoding proteins localized in mitochondria that are significantly deregulated (p<0.05). **(C)** The blue points indicate the genes encoding proteins reported deregulated with aging in literature that are significantly deregulated (p<0.05).

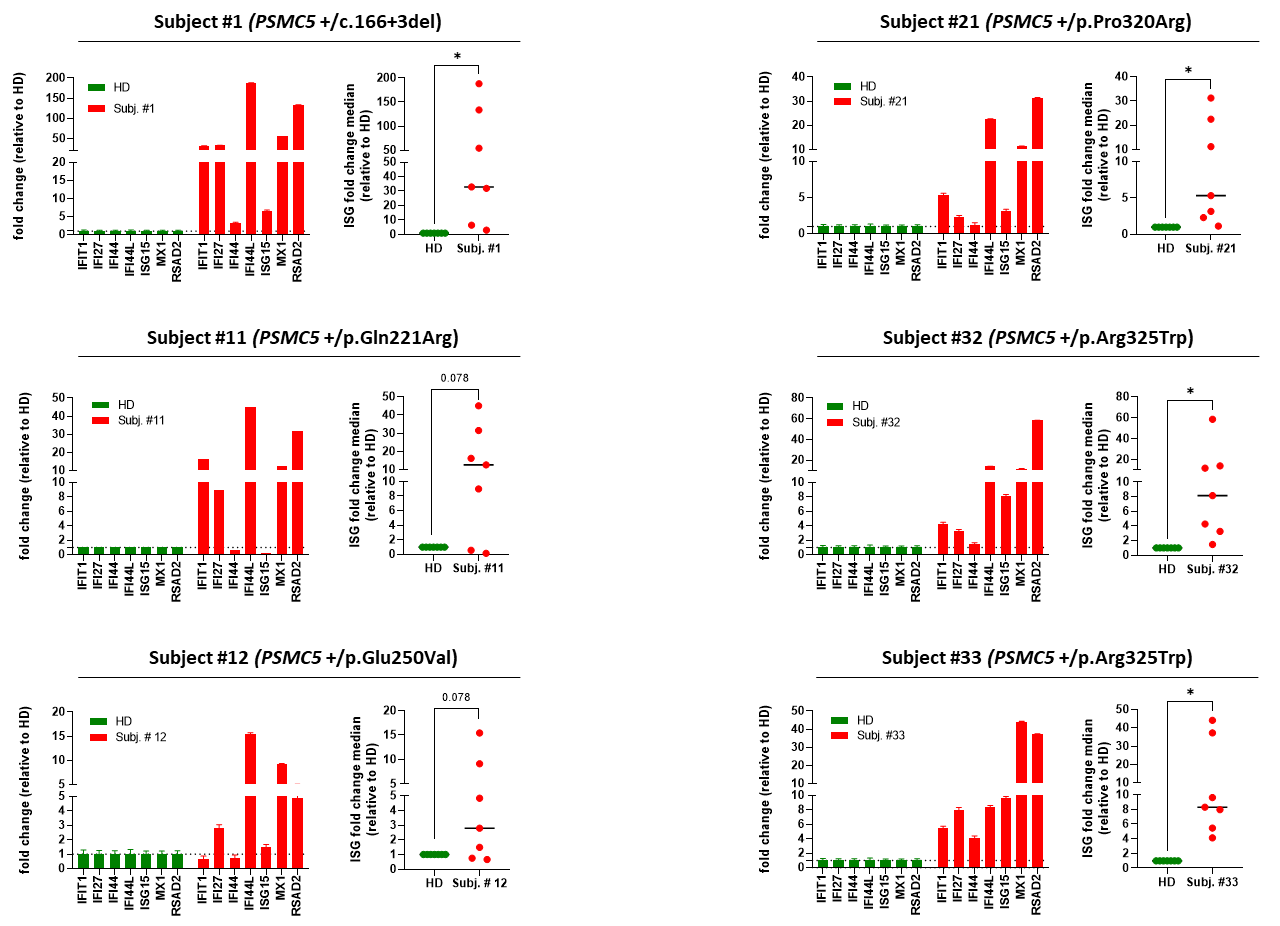

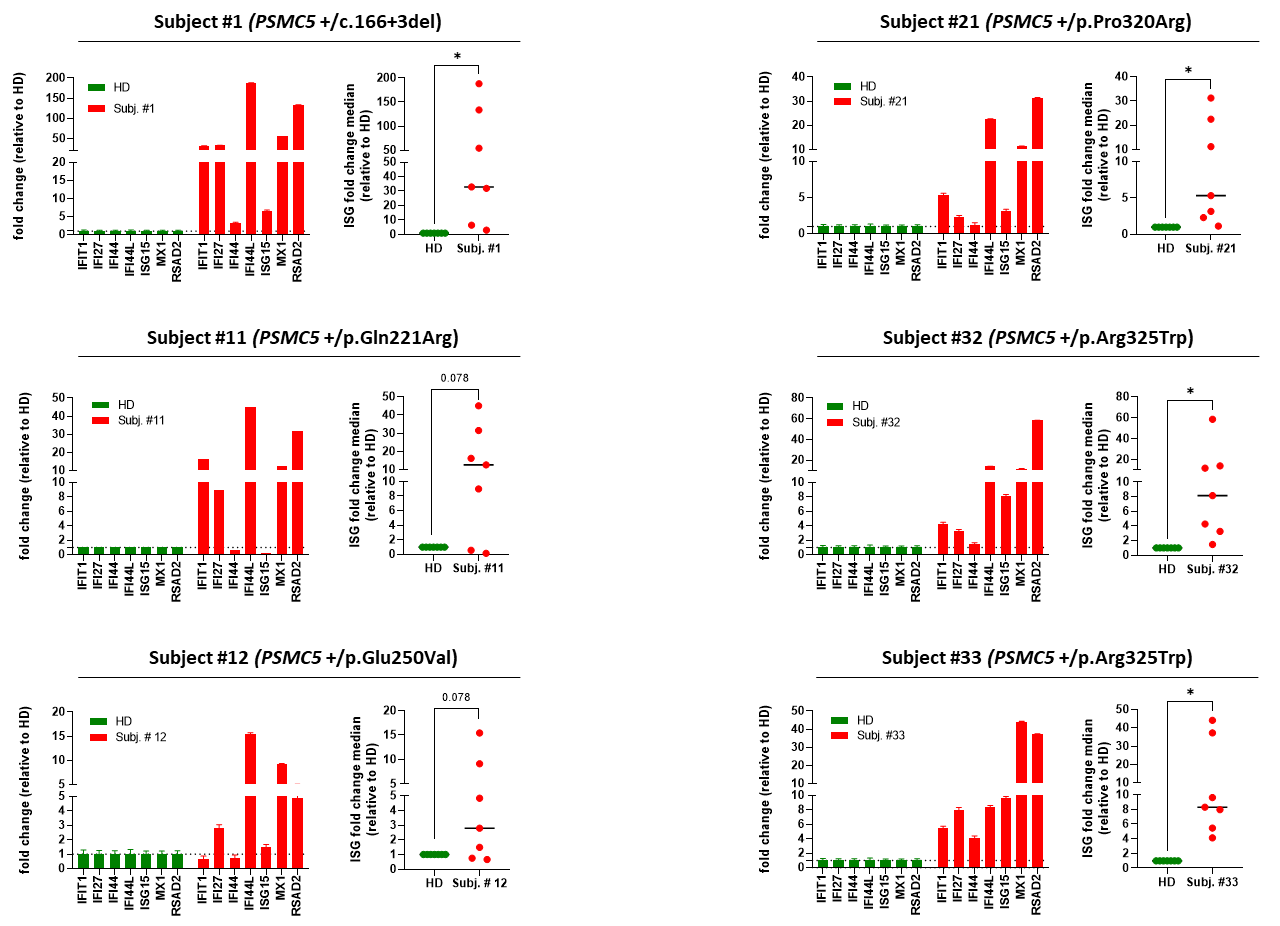

**Fig. S7: T cells from NDD subjects with *PSMC5* heterozygous variants exhibit a strong upregulation of ISG.** Gene expression of seven typical ISG (*IFIT1*, *IFI27*, *IFI44*, *IFI44L*, *ISG15*, *MX1* and *RSAD2*) was assayed by RT-qPCR on T cells derived from NDD subjects S1, S11, S12, S21, S32 and S33 as well as from healthy unrelated donors (HD), as indicated. Expression levels were normalized to GAPDH and relative quantifications (RQ) are presented as fold change for each ISG (left) or fold change median for all ISG (right) over control T cells (*p<0.05, Wilcoxon matched pairs signed rank test).

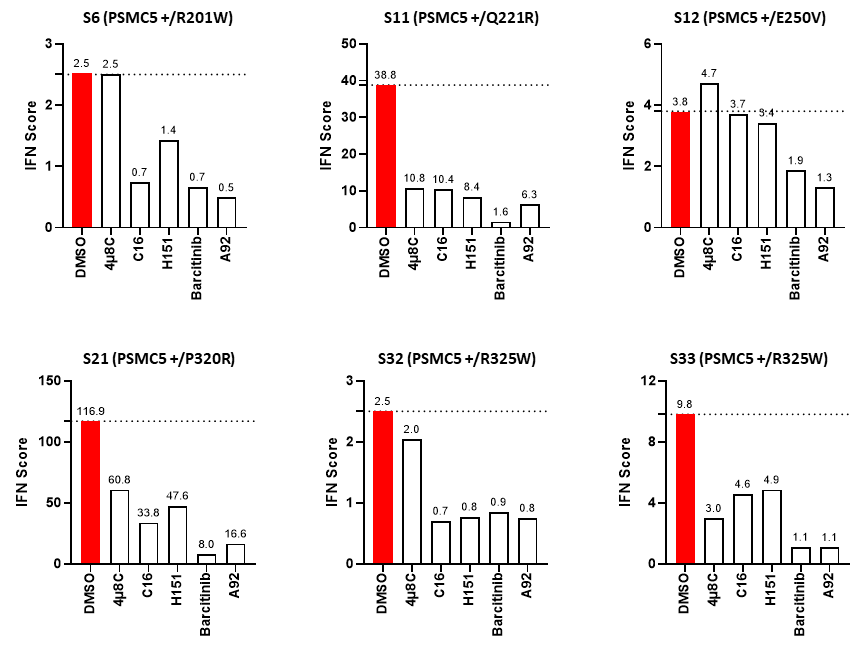

**Fig. S8: Sterile type I interferon (IFN) responses triggered in NDD patients with *PSMC5* heterozygous variants predominantly rely on the ISR**. T cells expanded from PBMC isolated from NDD subjects S6, S11, S12, S21, S32 and S33 were subjected to an 8-h treatment with DMSO (vehicle, marked in red), 4µ8C (100 µM), C16 (3 µM), H-151 (2 µM), baricitinib (1 µM) or A92 (10 µM) prior to RNA extraction and RT-qPCR for expression analysis of *IFIT1*, *IFI27*, *IFI44*, *IFI44L*, *ISG15*, *MX1* and *RSAD2*. Shown are the IFN scores calculated for each T cell sample under different treatment conditions.
